# Metabolome-wide Association Study of Bronchopulmonary Dysplasia in Extremely Premature Infants

**DOI:** 10.64898/2026.11.06.26367452

**Authors:** Thaybeth I. Malave-Mendez, Miguel Guardado, Cheryl Chapin, Ryan D. Hernandez, Philip L. Ballard, Dara G. Torgerson

## Abstract

**Rationale:** Untargeted metabolomics may identify novel pathways involved in bronchopulmonary dysplasia (BPD), a poorly understood lung disease in extremely premature infants.

**Objective:** To identify metabolites and pathways altered in infants who develop BPD through a metabolome-wide association study.

**Methods:** We performed untargeted urinary metabolomics in 314 infants (<29 weeks gestational age) at two time points between postnatal days 5-15 and 23-32, with detailed time-series for 28 infants. We compared metabolite levels between infants with and without BPD at 36 weeks postmenstrual age using logistic regression and pathway enrichment analysis. A stratified analysis by nutrition type was performed at 23-32 days.

**Results:** Among the 913 analyzed urinary metabolites, 23 and 68 were nominally associated with BPD between postnatal days 5-15 and 23-32, respectively (*p*<0.05). We found enrichment in the lactoyl amino acid sub-pathway (5-15 days; *p*=0.005, *q*=0.4) and xanthine metabolism and fatty acid metabolism (23-32 days; *p≤*0.002, *q*<0.1). Most BPD-differential metabolites were lower in infants with BPD at days 23-32 (OR=7.1, *p*=2×10^-6^), with similar patterns among infants receiving parenteral (OR=9.2, *p*=7×10^-7^) and full enteral nutrition (OR=6.1, *p=*0.003). Twenty-four metabolites, including vitamins C, B_1_, and B_6_, were lower in infants on parenteral nutrition with BPD at postnatal days 23-32.

**Discussion:** BPD in extremely premature infants was associated with lower levels of vitamin-related metabolites and dysregulation of lactoyl amino acids, xanthine, and fatty acid metabolism. BPD-differential metabolites were lower in infants with BPD during the fourth week of life, potentially related to higher energy demand, metabolic dysregulation, insufficient nutrient availability, and/or impaired intestinal absorption.

**What is already known on this topic:** Metabolic dysregulation has been implicated in bronchopulmonary dysplasia (BPD), but the metabolic pathways associated with BPD across early postnatal development remain poorly characterized. Existing metabolomic studies have been limited by small sample sizes and differences in biofluids, metabolomic approaches, and timing of sample collection, and no established metabolic biomarker exists for early diagnosis or monitoring of disease progression.

**What this study adds:** In a large, two-cohort metabolome-wide study of extremely premature infants, BPD was associated with distinct metabolic patterns that varied by postnatal age and nutrition source, with enrichment of xanthine and fatty acid metabolism during the fourth week of life. BPD-differential metabolites were predominantly lower during the fourth week of life, particularly among infants still receiving parenteral nutrition.

**How this study might affect research, practice or policy:** These findings identify metabolic and nutritional pathways for further investigation and may inform future studies of nutritional interventions aimed at reducing BPD risk in extremely premature infants.

## INTRODUCTION

Bronchopulmonary dysplasia (BPD) is a developmental lung disease and major contributor to neurodevelopmental and growth abnormalities, long-term respiratory morbidity, and mortality among extremely low gestational age newborns [1,2]. Heterogeneity of BPD and advances in neonatal healthcare have led to evolving definitions [3]. The recently described “new BPD” in extremely premature infants is characterized by impaired alveolar development and dysregulated pulmonary vasculature, leading to persistent abnormalities in gas exchange, airway function, respiratory mechanics, and lung volumes [4]. Risk factors associated with BPD include low gestational age and birthweight, male sex, and social and environmental factors that sometimes correlate with race/ethnicity [5,6]. Despite advances in neonatal healthcare, there has been minimal change in the incidence of BPD during the last 20 years [7]. The limited understanding of BPD pathophysiology has hindered the development of effective diagnostics, preventive therapies, and treatments that may help lower the incidence of BPD [7,8]. Novel approaches are needed to identify the biological mechanisms involved in BPD.

Metabolomics involves the quantification of many hundreds of biochemicals (metabolites) that are present in a biological sample and has emerged as a powerful approach for studies of complex disease. It can simultaneously capture the combined effects of environmental and genetic factors that may impact human physiology, and increase our overall comprehension of disease pathogenesis [9]. Previous studies have implicated metabolic dysregulation in amino acid, lipid, and/or carnitine pathways in BPD; however, there is a limited consensus across studies [10–20]. This may be due to varying infant demographics, small sample sizes of individual studies (<100 preterm infants), differences in the type of biofluid sampled, and reliance on single time point sampling during the clinical course of premature infants in the neonatal intensive care unit (NICU). Furthermore, the majority of studies have used a targeted approach to characterize metabolites of interest, whereas untargeted metabolomics can query a greater number of metabolites to test for associations with disease and provide a broader view of the pathways involved.

In this study, we characterized the urinary metabolome of 314 extremely premature infants at high risk of developing BPD (including 178 infants diagnosed with BPD at 36 weeks) using an untargeted approach to identify a broad array of metabolites present in urine. We performed a metabolome-wide association study (MWAS) for BPD at 36 weeks postmenstrual age to identify metabolites and metabolic pathways associated with BPD, including multiple time point assessments in the first 30 days of life. Our results provide insight into the metabolic pathways associated with BPD development and offer information to support future studies of early diagnosis and the development of more effective therapies and interventions. Some of the results of this study have been previously reported in abstract form [21].

## METHODS

### Study Description

A total of 171 infants from the Trial of Late Surfactant for Prevention of Bronchopulmonary Dysplasia (TOLSURF, NCT01022580) and 143 infants from the Prematurity and Respiratory Outcomes Program (PROP, NCT01435187) were included in our study (details in data supplement). Clinical data were obtained for both cohorts and summarized in Table 1. BPD was assessed at 36 weeks postmenstrual age by physiological testing [22]. Research protocols were approved by the institutional review boards of the participating institutions, and a parent of each infant provided written informed consent.

**Table 1.** Characteristics of infants from TOLSURF and PROP included in metabolomic studies by BPD status at 36 weeks postmenstrual age.

|  | TOLSURF (n = 171) |  |  | PROP (n = 143) |  |  |
|---|---|---|---|---|---|---|
|  | BPD | No BPD | *p* | BPD | No BPD | *p* |
| Number of Infants | 106 | 65 | - | 72 | 71 | - |
| Number of Infants by Maternal Race/Ethnicity<sup>†</sup> |  |  | 0.011 |  |  | 1 |
| *Black or African American* | 30 (28) | 31 (48) |  | 30 (42) | 30 (42) |  |
| *Hispanic* | 14 (13) | 11 (17) |  | 12 (16) | 11 (16) |  |
| *White* | 62 (59) | 23 (35) |  | 30 (42) | 30 (42) |  |
| Gestational Age in weeks<sup>†</sup> | 25.3 (1.3) | 25.4 (1.1) | 0.58 | 25.9 (1.1) | 25.8 (1.3) | 0.42 |
| Sex (male)<sup>†</sup> | 64 (60.4) | 36 (55.4) | 0.53 | 41 (56.9) | 34 (47.9) | 0.32 |
| Birthweight (g)<sup>†</sup> | 706 (172) | 757 (177) | 0.064 | 766 (135) | 805 (163) | 0.12 |
| Average RSS days 7-14 | 4.5 (2.1) | 3.5 (1.3) | 1.2x10<sup>-4</sup> | 2.8 (1.7) | 2.4 (1.6) | 0.077 |
| Average RSS days 21-28 | 4.4 (2.3) | 3.7 (2.1) | 1.8x10<sup>-15</sup> | 4.1 (3.0) | 1.7 (1.3) | 2.8x10<sup>-15</sup> |
| Age days sample 1 | 9.4 (2.4) | 9.8 (2.3) | 0.35 | 6.6 (1.1) | 6.8 (1.0) | 0.40 |
| Age days sample 2 | 27.5 (2.1) | 27.7 (2.3) | 0.68 | 28.0 (1.0) | 28.3 (0.86) | 0.13 |
| BPD at 40 weeks | 69 (65.1) | 0 (0) | - | 46 (63.0) | 1 (1.4) | - |
| On PN sample 1 | 106 (100) | 65 (100) | 1 | 72 (100) | 71 (100) | 1 |
| On PN sample 2<sup>*</sup> | 67 (63.2) | 42 (64.6) | 0.87 | 44 (61.1) | 51 (71.8) | 0.22 |
| On corticosteroids sample 1<sup>†</sup> | 8 (7.5) | 6 (9.2) | 0.78 | 3 (4.2) | 1 (1.4) | 0.62 |
| On corticosteroids sample 2<sup>†</sup> | 22 (20.8) | 14 (21.5) | 1 | 20 (27.8) | 21 (29.6) | 0.85 |
| On caffeine sample 1 | 83 (78.3) | 55 (84.6) | 0.42 | 51 (70.8) | 53 (74.6) | 0.71 |
| On caffeine sample 2 | 90 (84.9) | 60 (92.3) | 0.22 | 60 (83.3) | 65 (91.5) | 0.21 |

*TOLSURF* Trial of Late Surfactant for Prevention of Bronchopulmonary Dysplasia; *PROP* Prematurity and Respiratory Outcomes Program; *BPD* Bronchopulmonary Dysplasia; *p* p-value; *RSS* Respiratory Severity Score; *PN* Parenteral Nutrition
Values are represented by the mean (standard deviation) for continuous variables and the count (percent) for categorical variables. Quantitative measurements were compared using a t-test, and categorical data were compared using a Fisher’s exact test.
<sup>†</sup>Covariates included in the MWAS.
\*Additional covariate included in the MWAS at time point 2.

### Sample Selection

Urine samples were collected from each infant and stored at - 80°C until processing. Infants were selected based on survival at 40 weeks gestational age and availability of urine samples. Two samples were selected for each infant, one collected between postnatal days 5-15 (time point 1, mean±SD 8±2 days) and another between postnatal days 23-32 (time point 2, mean±SD 28±2 days).

### Metabolomics Profiling

Untargeted metabolomic assays were performed using Ultra High-Performance Liquid Chromatography-Tandem Mass Spectroscopy (UHPLC/MS-MS) by Metabolon Inc. as previously described [23,24]. Metabolic profiling was conducted separately for each cohort, with anchor samples included to facilitate the comparison of metabolite abundance between cohorts. Metabolite levels were normalized to osmolality and median scaled. Metabolites with >30% of values below the limit of detection within either cohort were removed from further analysis. A detailed description of metabolomic profiling is provided in the data supplement.

### Statistical Analyses

Principal component analysis (PCA) was performed across all metabolites to evaluate global differences in the urinary metabolome. A Mann-Whitney *U*-test was performed to assess differences in the first two principal components (PCs) for infants with and without BPD. At time point 2, PCA was also performed separately by nutrition source (parenteral vs full enteral nutrition).

A metabolome-wide association study (MWAS) was performed using logistic regression to test for associations between BPD and quantitative levels of individual metabolites at each time point and the fold-change in levels at time point 2 vs time point 1. Covariates included clinical and demographic characteristics (Table 1). Analyses were done within each cohort and combined using a fixed-effects meta-analysis. Sub-analyses were performed by nutrition type at time point 2 to evaluate whether patterns of association with BPD differed by nutrition source. Details on infant feeding status categorization, statistical models, and covariates are included in the data supplement.

Metabolites with nominal *p*<0.05 were carried forward for exploratory characterization and pathway enrichment analyses and are referred to as “BPD-differential metabolites” throughout. A Fisher’s exact test was used to assess whether BPD-differential metabolites were more likely than non-differential metabolites to have lower versus higher levels in infants with BPD, with odds ratios used to quantify the directional enrichment. To account for multiple testing, false discovery rates were estimated separately within each MWAS using the qvalue package in R [25]. Metabolites of known identity were annotated to 88 sub-pathways, and 91 sub-pathways at time points 1 and 2, respectively (Metabolon Inc, April 2025). Pathway enrichment analysis was performed for each super-pathway and sub-pathway using a Fisher’s exact test for each MWAS comparison (additional details in data supplement).

Longitudinal data for selected BPD-differential metabolites from a prior study [24] were aligned to days relative to discontinuation of parenteral nutrition and visualized using time-course plots with locally estimated scatterplot smoothing (LOESS) by BPD status. Data included untargeted metabolomics of 302 urine samples from 28 infants (17 with BPD and 11 without BPD) in TOLSURF (additional details in data supplement), of whom 25 were also included in the current two-time point study.

Analyses were performed using Python 3.11.4 [26] and R 4.5.1 [27].

## RESULTS

Clinical and demographic characteristics were similar between infants with and without BPD at 36 weeks postmenstrual age, apart from infants with BPD having higher respiratory severity scores (RSS) at both time points (Table 1). Mean values across groups for gestational age ranged between 25-26 weeks, and birthweight between 720-790 g. Untargeted urinary metabolomic profiling identified 1441 unique metabolites across both cohorts at time points 1 and 2, of which 874 and 913 passed quality control thresholds, respectively (851 overlapping between time points). The first two principal components across all metabolites showed little to no separation of infants by BPD status (*p*>0.05; Figure 1A-D; Figure E1A-B, E1D), apart from infants on parenteral nutrition at time point 2 (PC1; *p*=0.042; Figure E1C).

**Figure 1.**
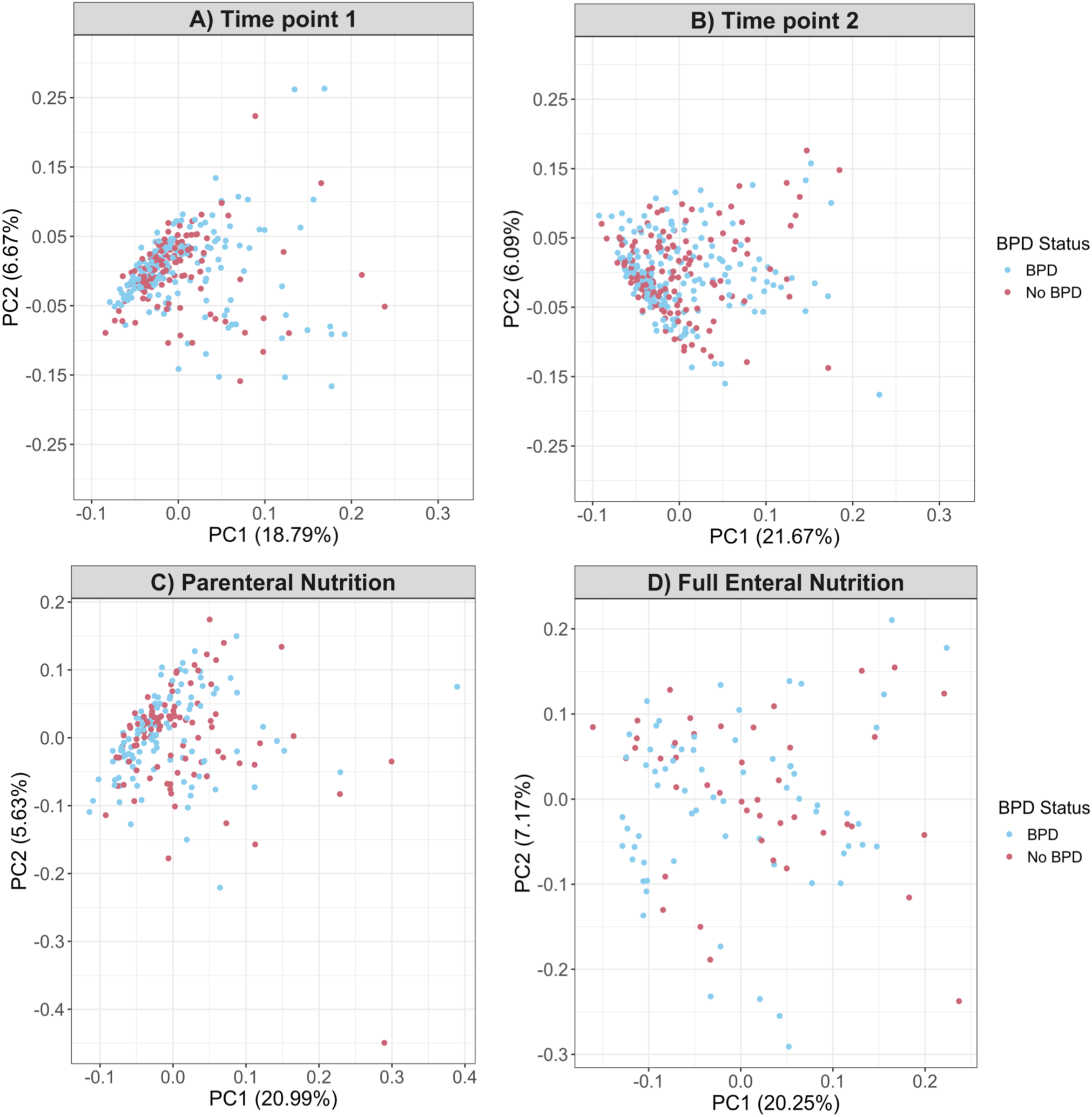
Principal component analyses (PCA) of urinary metabolite levels of infants from both cohorts at two sampling time points. Scatter plots of the first two principal components (PC1 and PC2). (A) PCA of 874 biochemicals detected in urine samples collected between postnatal days 5-15 (time point 1) across all infants. (B) PCA of 913 biochemicals detected in urine samples collected between postnatal days 23-32 (time point 2) across all infants. At time point 2, PCA was performed separately among infants receiving (C) parenteral nutrition and (D) full enteral nutrition. Percent values on each axis indicate the variance explained.

### Metabolome-Wide Association Study (MWAS)

We tested for an association between BPD and levels of urinary metabolites for 1) each time point separately, 2) fold-change across time points, and 3) sub-analyses at time point 2 with infants separated by feeding modality (parenteral vs. full enteral).

Levels of 23 and 68 individual metabolites were nominally associated with BPD (*p*<0.05) at time points 1 and 2, respectively (Figure 2A-B and Supplemental Table E1-E2), and 28 metabolites were nominally associated with BPD based on fold-change between time points (Table E3). Odds ratios for the 10 BPD-differential metabolites with the smallest *p*-values are shown in Figure 3, with cohort-specific effect estimates in the same direction. Across BPD-differential metabolites, 13 and 44 were of known identity at time points 1 and 2, respectively, and belonged to a variety of sub-pathways (Table 2, Table 3). Five BPD-differential metabolites were identified at both time points, including 3-hydroxykynurenine (tryptophan metabolism), pipecolate (lysine metabolism), beta-hydroxyisovaleroylcarnitine (leucine/isoleucine/valine metabolism), and two unnamed metabolites. One of these, 3-hydroxykynurenine, showed an opposite direction of association with BPD across time points, with higher levels in infants with BPD at time point 1 and lower levels at time point 2. A similar trend was observed in longitudinal data following discontinuation of parenteral nutrition (Figure 4A). Pipecolate was higher in infants with BPD at both time points, and beta-hydroxyisovaleroylcarnitine and two unnamed metabolites were lower in infants with BPD at both time points. Fold-change over time in 19 metabolites of known identity was nominally associated with BPD, including hexanoylcarnitine (C6) and four unnamed metabolites that were also BPD-differential at time point 2.

**Figure 2.**
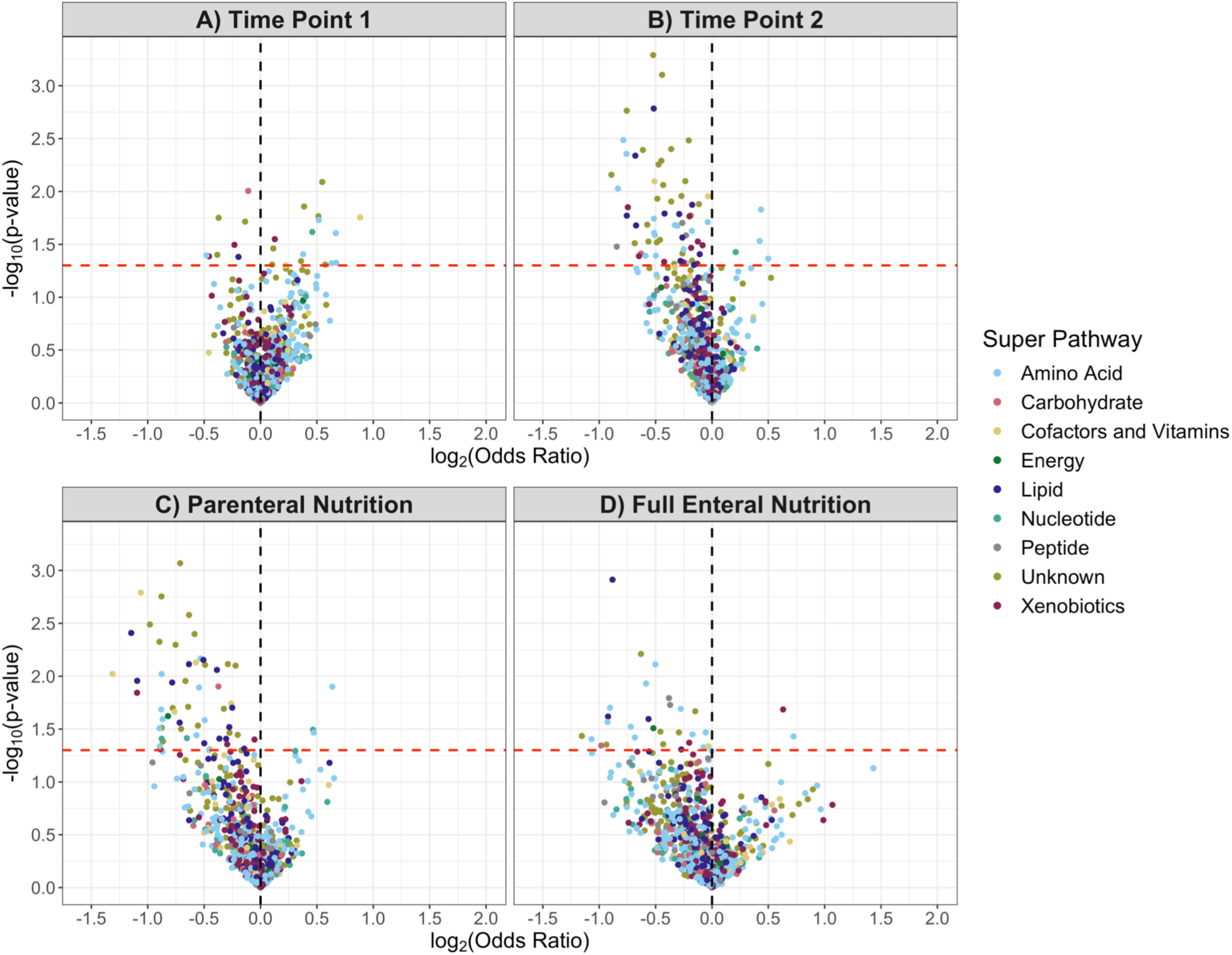
Volcano plot identifying BPD-differential metabolites at two sampling time points. Metabolite odds ratios are presented as log_2_(OR), and nominal *p*-values as - log10(*p*-value). The dashed red line indicates the nominal *p*=0.05 threshold used to define BPD-differential metabolites, and the dashed black line divides metabolites higher in infants who developed BPD (log_2_(OR)>0) from those higher in infants who did not (log_2_(OR)<0). Each metabolite is represented by a dot colored according to its super pathway. (A) Days 5-15 postnatal (time point 1) across all infants, (B) Days 23-32 postnatal (time point 2) across all infants, (C) Time point 2 among infants on parenteral nutrition, (D) Time point 2 among infants on full enteral nutrition.

**Figure 3.**
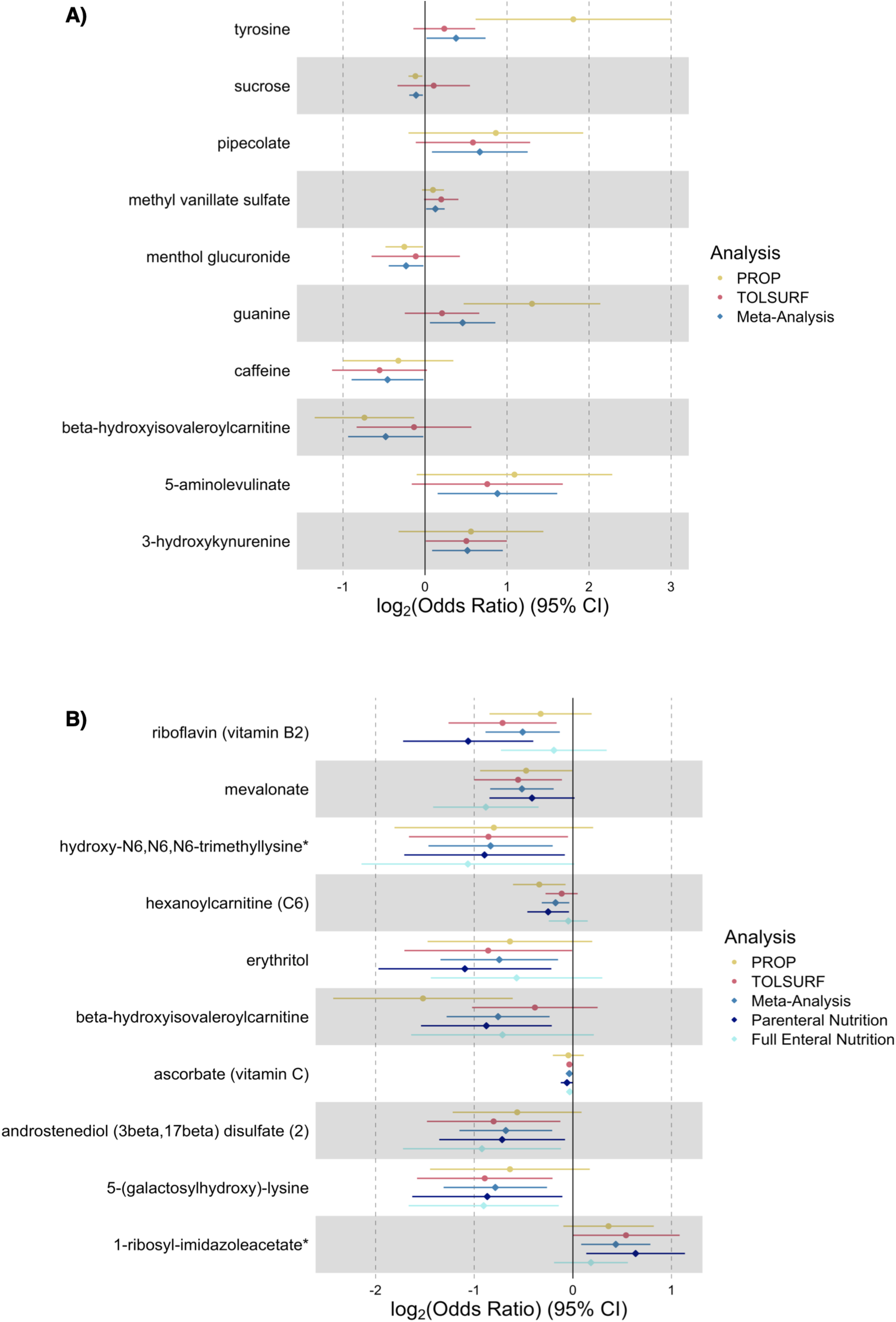
Odds ratios for the 10 BPD-differential metabolites with the smallest meta-analysis *p*-values. Metabolites are shown in ascending *p*-value order from top to bottom. Horizontal error bars denote 95% confidence intervals. The solid vertical black line at log_2_(OR)=0 separates estimates corresponding to higher metabolite levels in infants who developed BPD (log_2_(OR)>0) from those corresponding to higher levels in infants who did not (log_2_(OR)<0). Odds ratio estimates are shown for the PROP cohort in yellow, the TOLSURF cohort in red, and the meta-analysis in blue at (A) time point 1 and (B) time point 2. Time point 2 also includes odds ratios from the nutrition-stratified analyses, represented by dark blue for infants on parenteral nutrition and light blue for those on full enteral nutrition.

**Figure 4.**
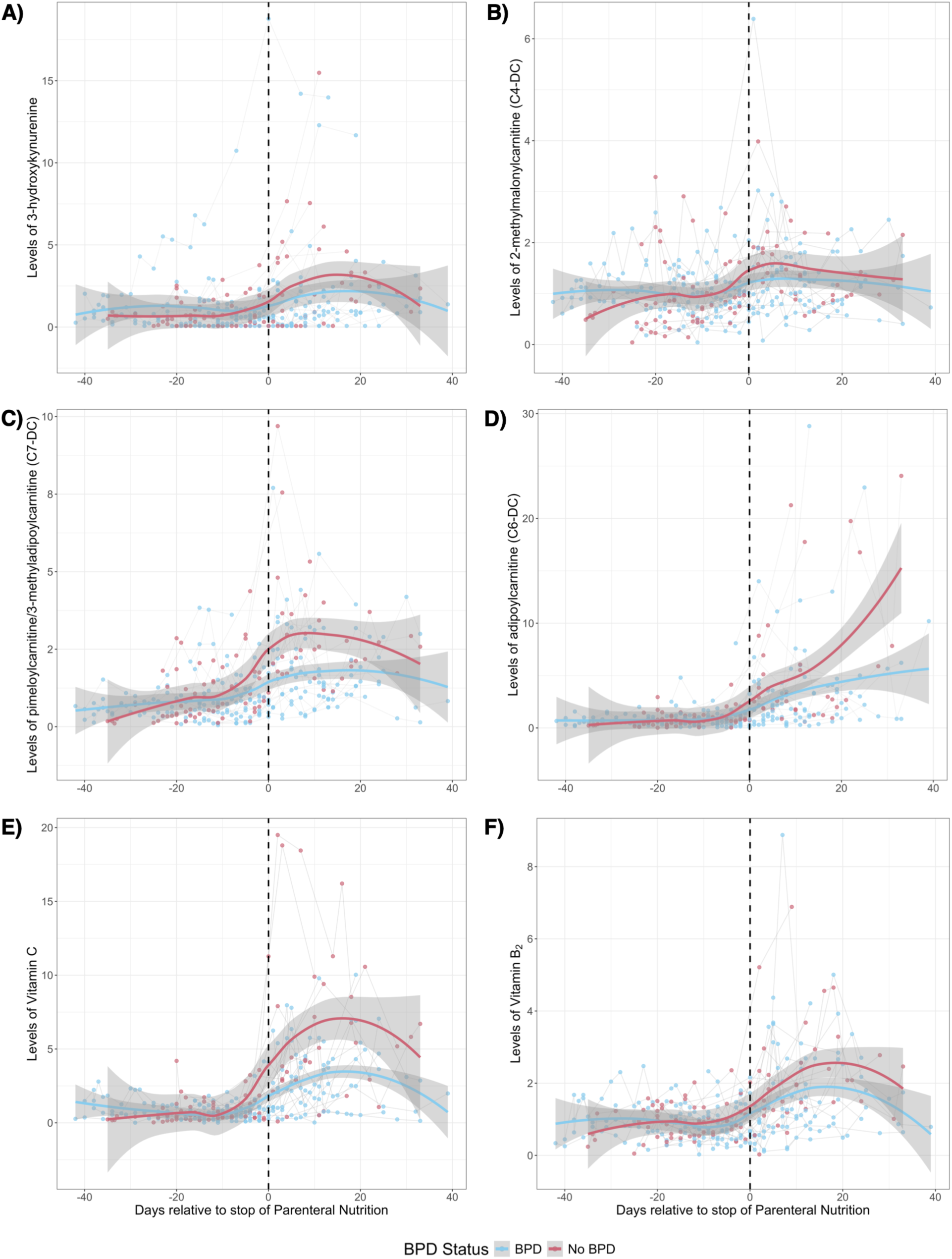
Longitudinal trajectories of selected individual metabolites relative to parenteral nutrition discontinuation by BPD status. Locally estimated scatterplot smoothing (LOESS) plots are shown for (A) 3-hydroxykynurenine, (B) 2-methylmalonylcarnitine (C4-DC), (C) pimeloylcarnitine/3-methyladipoylcarnitine (C7-DC), (D) adipoylcarnitine (C6-DC), (E) vitamin C (ascorbic acid 3-sulfate*), and (F) vitamin B_2_ (riboflavin). Points represent individual observations, with connected points indicating measurements from the same infant. Shaded bands represent 95% confidence intervals around the LOESS fit for descriptive visualization and do not account for within-infant correlation from repeated measurements. The dashed vertical line indicates the day parenteral nutrition was discontinued (day 0). For all metabolites shown, increasing levels are generally observed after stopping parenteral nutrition, with visually slower increases among infants who develop BPD.

**Table 2.** BPD-differential metabolites of known identity during postnatal days 5-15 (time point 1).

| Metabolite Name | Sub-pathway | TOLSURF |  | PROP |  | Meta |  |  |
|---|---|---|---|---|---|---|---|---|
|  |  | Beta (SE) | *p* | Beta (SE) | *p* | Beta (SE) | *p* | *q* |
| sucrose | Disaccharides and Oligosaccharides | 0.074 (0.156) | 0.638 | -0.081 (0.030) | 0.007 | -0.076 (0.029) | 0.010 | 0.998 |
| 4-methylnonanoylcarnitine | Fatty Acid Metabolism (Acyl Carnitine, Medium Chain) | 0.021 (0.104) | 0.838 | -0.244 (0.087) | 0.005 | -0.136 (0.067) | 0.042 | 0.998 |
| methyl vanillate sulfate | Food Component/Plant | 0.137 (0.074) | 0.064 | 0.067 (0.048) | 0.155 | 0.088 (0.040) | 0.028 | 0.998 |
| menthol glucuronide |  | -0.079 (0.191) | 0.680 | -0.175 (0.081) | 0.031 | -0.160 (0.075) | 0.032 | 0.998 |
| 5-aminolevulinate | Hemoglobin and Porphyrin Metabolism | 0.526 (0.326) | 0.106 | 0.756 (0.422) | 0.073 | 0.612 (0.258) | 0.018 | 0.998 |
| N-lactoyl isoleucine | Lactoyl Amino Acid | 0.389 (0.265) | 0.143 | 0.546 (0.398) | 0.171 | 0.437 (0.221) | 0.048 | 0.998 |
| N-lactoyl leucine |  | 0.469 (0.290) | 0.106 | 0.458 (0.398) | 0.250 | 0.465 (0.234) | 0.047 | 0.998 |
| beta-hydroxyisovaleroylcarnitine | Leucine, Isoleucine and Valine Metabolism | -0.093 (0.248) | 0.707 | -0.512 (0.214) | 0.017 | -0.332 (0.162) | 0.040 | 0.998 |
| pipecolate | Lysine Metabolism | 0.405 (0.247) | 0.100 | 0.599 (0.377) | 0.112 | 0.463 (0.206) | 0.025 | 0.998 |
| guanine | Purine Metabolism, Guanine containing | 0.144 (0.161) | 0.371 | 0.904 (0.295) | 0.002 | 0.318 (0.141) | 0.024 | 0.998 |
| 3-hydroxykynurenine | Tryptophan Metabolism | 0.349 (0.175) | 0.045 | 0.388 (0.312) | 0.214 | 0.359 (0.152) | 0.019 | 0.998 |
| tyrosine | Tyrosine Metabolism | 0.163 (0.133) | 0.222 | 1.253 (0.421) | 0.003 | 0.262 (0.127) | 0.039 | 0.998 |
| caffeine | Xanthine Metabolism | -0.385 (0.205) | 0.060 | -0.226 (0.237) | 0.342 | -0.317 (0.155) | 0.041 | 0.998 |

*Meta* meta-analysis; *Beta* beta-coefficient; *SE* standard error; *p* p-value; *q* q-value representing the false discovery rate associated with each metabolite
Analysis used logistic regression for the TOLSURF and PROP cohorts separately, adjusting for gestational age, birthweight, sex, self-reported maternal race/ethnicity, and corticosteroids (>1 day), followed by a meta-analysis. A negative effect size/unit change (beta) indicates metabolites of lower abundance in BPD infants. Metabolites are grouped by sub-pathway. Sub-pathways and metabolites are listed alphabetically. Complete results are provided in the supplementary material.

**Table 3.** BPD-differential metabolites of known identity during postnatal days 23-32 (time point 2).

| Metabolite Name | Sub-pathway | TOLSURF |  | PROP |  | Meta |  |  |
|---|---|---|---|---|---|---|---|---|
|  |  | Beta (SE) | *p* | Beta (SE) | *p* | Beta (SE) | *p* | *q* |
| N-acetylglucosaminylasparagine | Aminosugar Metabolism | -0.470 (0.271) | 0.084 | -0.385 (0.335) | 0.251 | -0.436 (0.211) | 0.039 | 0.632 |
| androstenediol (3beta,17beta) disulfate (1) | Androgenic Steroids | -0.161 (0.134) | 0.227 | -0.231 (0.141) | 0.102 | -0.194 (0.097) | 0.045 | 0.658 |
| androstenediol (3beta,17beta) disulfate (2) |  | -0.557 (0.239) | 0.02 | -0.391 (0.231) | 0.091 | -0.471 (0.166) | 0.005 | 0.419 |
| **ascorbate (vitamin C)** | Ascorbate and Aldarate Metabolism | -0.025 (0.010) | 0.013 | -0.032 (0.056) | 0.571 | -0.025 (0.010) | 0.011 | 0.520 |
| diaminopimelate | Bacterial/Fungal | -0.194 (0.130) | 0.134 | -0.159 (0.120) | 0.186 | -0.175 (0.088) | 0.047 | 0.658 |
| sulfate* | Chemical | -0.533 (0.301) | 0.077 | -0.354 (0.321) | 0.271 | -0.449 (0.220) | 0.041 | 0.611 |
| 3-hydroxypyridine sulfate |  | -0.113 (0.071) | 0.113 | -0.168 (0.115) | 0.143 | -0.128 (0.061) | 0.034 | 0.632 |
| phenylalanylhydroxyproline* | Dipeptide | -0.286 (0.111) | 0.01 | -0.079 (0.111) | 0.479 | -0.183 (0.078) | 0.02 | 0.520 |
| leucylhydroxyproline* | Dipeptide Derivative | -0.205 (0.092) | 0.026 | -0.089 (0.116) | 0.443 | -0.160 (0.072) | 0.026 | 0.579 |
| pimeloylcarnitine/3-methyladipoylcarnitine (C7-DC) | Fatty Acid Metabolism (Acyl Carnitine, Dicarboxylate) | -0.206 (0.120) | 0.086 | -0.197 (0.117) | 0.093 | -0.201 (0.084) | 0.016 | 0.520 |
| 3-hydroxyoctanoylcarnitine (1) | Fatty Acid Metabolism (Acyl Carnitine, Hydroxy) | -0.053 (0.101) | 0.604 | -0.440 (0.143) | 0.002 | -0.182 (0.083) | 0.027 | 0.587 |
| hexanoylcarnitine (C6) | Fatty Acid Metabolism (Acyl Carnitine, Medium Chain) | -0.079 (0.058) | 0.173 | -0.236 (0.094) | 0.012 | -0.122 (0.049) | 0.013 | 0.520 |
| octanoylcarnitine (C8) |  | -0.054 (0.037) | 0.143 | -0.861 (0.226) | 1.4x10<sup>-4</sup> | -0.075 (0.037) | 0.039 | 0.632 |
| 2-methylhexanoylcarnitine |  | -0.056 (0.061) | 0.362 | -0.428 (0.147) | 0.004 | -0.111 (0.056) | 0.049 | 0.658 |
| 2-methylmalonylcarnitine (C4-DC) | Fatty Acid Metabolism (also BCAA Metabolism) | -0.280 (0.242) | 0.247 | -0.902 (0.369) | 0.014 | -0.467 (0.202) | 0.021 | 0.520 |
| 2-hydroxyadipate | Fatty Acid, Dicarboxylate | -0.447 (0.277) | 0.107 | -0.653 (0.359) | 0.069 | -0.524 (0.219) | 0.017 | 0.520 |
| erythritol | Food Component/Plant | -0.595 (0.301) | 0.048 | -0.442 (0.295) | 0.134 | -0.517 (0.211) | 0.014 | 0.520 |
| 1-methylguanidine | Guanidino and Acetamido Metabolism | -0.128 (0.120) | 0.285 | -0.439 (0.174) | 0.012 | -0.228 (0.099) | 0.021 | 0.520 |
| 4-imidazoleacetate | Histidine Metabolism | 0.426 (0.265) | 0.108 | 0.289 (0.224) | 0.197 | 0.346 (0.171) | 0.043 | 0.520 |
| 1-ribosyl-imidazoleacetate* |  | 0.374 (0.192) | 0.052 | 0.251 (0.162) | 0.122 | 0.302 (0.124) | 0.015 | 0.520 |
| N-acetyl-1-methylhistidine* |  | -0.351 (0.182) | 0.053 | -0.369 (0.271) | 0.174 | -0.357 (0.151) | 0.018 | 0.656 |
| beta-hydroxyisovaleroyl carnitine | Leucine, Isoleucine and Valine Metabolism | -0.267 (0.225) | 0.236 | -1.053 (0.322) | 0.001 | -0.525 (0.185) | 0.004 | 0.419 |
| tiglylcarnitine (C5:1-DC) |  | -0.135 (0.184) | 0.463 | -0.980 (0.310) | 0.002 | -0.356 (0.158) | 0.025 | 0.520 |
| 3-methylglutarylcarnitine (2) |  | -0.078 (0.123) | 0.523 | -0.389 (0.140) | 0.006 | -0.213 (0.092) | 0.021 | 0.567 |
| methylsuccinoylcarnitine |  | -0.095 (0.082) | 0.246 | -0.147 (0.073) | 0.045 | -0.124 (0.055) | 0.024 | 0.576 |
| pipecolate | Lysine Metabolism | 0.395 (0.193) | 0.041 | 0.198 (0.188) | 0.293 | 0.294 (0.135) | 0.029 | 0.419 |
| 5-(galactosylhydroxy)-lysine |  | -0.619 (0.243) | 0.011 | -0.442 (0.286) | 0.123 | -0.545 (0.185) | 0.003 | 0.504 |
| hydroxy-N6,N6,N6-trimethyllysine* |  | -0.593 (0.285) | 0.038 | -0.555 (0.356) | 0.119 | -0.578 (0.223) | 0.009 | 0.587 |
| taurine | Methionine, Cysteine, SAM and Taurine Metabolism | -0.097 (0.114) | 0.396 | -0.209 (0.109) | 0.054 | -0.155 (0.079) | 0.048 | 0.658 |
| mevalonate | Mevalonate Metabolism | -0.385 (0.157) | 0.014 | -0.328 (0.165) | 0.046 | -0.358 (0.114) | 0.002 | 0.393 |
| N,N-dimethyl-pro-pro | Modified Peptides | -0.648 (0.374) | 0.083 | -0.511 (0.406) | 0.208 | -0.585 (0.275) | 0.033 | 0.607 |
| isoputreanine | Polyamine Metabolism | -0.308 (0.239) | 0.197 | -0.639 (0.362) | 0.078 | -0.408 (0.199) | 0.041 | 0.632 |
| 21-hydroxypregnenolone disulfate | Pregnenolone Steroids | -0.326 (0.180) | 0.07 | -0.263 (0.164) | 0.109 | -0.292 (0.121) | 0.016 | 0.52 |
| uridine | Pyrimidine Metabolism, Uracil containing | 0.101 (0.106) | 0.34 | 0.182 (0.094) | 0.053 | 0.146 (0.070) | 0.037 | 0.632 |
| **riboflavin (vitamin B2)** | Riboflavin Metabolism | -0.494 (0.194) | 0.011 | -0.227 (0.183) | 0.216 | -0.353 (0.133) | 0.008 | 0.489 |
| 5-hydroxyindoleacetate | Tryptophan Metabolism | -0.544 (0.233) | 0.02 | -0.129 (0.243) | 0.595 | -0.345 (0.168) | 0.04 | 0.619 |
| 3-hydroxykynurenine |  | -0.447 (0.198) | 0.024 | -0.116 (0.091) | 0.202 | -0.174 (0.083) | 0.035 | 0.632 |
| 4-hydroxyphenylacetoylcarnitine | Tyrosine Metabolism | -0.024 (0.014) | 0.081 | -0.032 (0.020) | 0.112 | -0.026 (0.011) | 0.019 | 0.520 |
| pro-hydroxy-pro | Urea cycle; Arginine and Proline Metabolism | -0.660 (0.269) | 0.014 | -0.190 (0.194) | 0.328 | -0.351 (0.157) | 0.026 | 0.579 |
| 3-methylxanthine | Xanthine Metabolism | -0.050 (0.043) | 0.24 | -0.063 (0.035) | 0.071 | -0.058 (0.027) | 0.032 | 0.520 |
| 7-methylxanthine |  | -0.061 (0.057) | 0.281 | -0.145 (0.077) | 0.061 | -0.090 (0.046) | 0.048 | 0.587 |
| 1,7-dimethylurate |  | -0.158 (0.090) | 0.079 | -0.124 (0.076) | 0.102 | -0.138 (0.058) | 0.017 | 0.601 |
| 1-methylurate |  | -0.496 (0.241) | 0.04 | -0.173 (0.186) | 0.354 | -0.294 (0.147) | 0.046 | 0.658 |
| 7-methylurate |  | -0.058 (0.052) | 0.267 | -0.129 (0.063) | 0.039 | -0.087 (0.040) | 0.03 | 0.658 |

*Meta* meta-analysis; *Beta* beta-coefficient; *SE* standard error; *p* p-value; *q* q-value representing the false discovery rate associated with each metabolite
Vitamins are in **boldface**.
Analysis used logistic regression for the TOLSURF and PROP cohorts separately, adjusting for gestational age, birthweight, sex, self-reported maternal race/ethnicity, nutrition source (parenteral vs enteral nutrition), and corticosteroids (>1 day), followed by a meta-analysis. A negative effect size/unit change (beta) indicates metabolites of lower abundance in BPD infants. Metabolites are grouped by sub-pathway. Sub-pathways and metabolites are listed alphabetically. Complete results are provided in the supplementary material.

Sub-analyses among infants separated by mode of nutrition at time point 2 identified 60 BPD-differential metabolites in those receiving parenteral nutrition and 30 in those receiving full enteral nutrition (Figure 2C-D; Supplemental Tables E4-E5), of which three overlapped in the same direction of effect [androstenediol (3beta,17beta) disulfate (2), 5-(galactosylhydroxy)-lysine, and an unnamed metabolite]. A total of 28 BPD-differential metabolites were unique to infants receiving parenteral nutrition (Table 4), and 13 were unique to infants on full enteral nutrition (Table 5). Of the 68 BPD-differential metabolites at time point 2 across all infants, 29 were identified among infants receiving parenteral nutrition and 14 among infants receiving full enteral nutrition (Figure 5).

**Figure 5.**
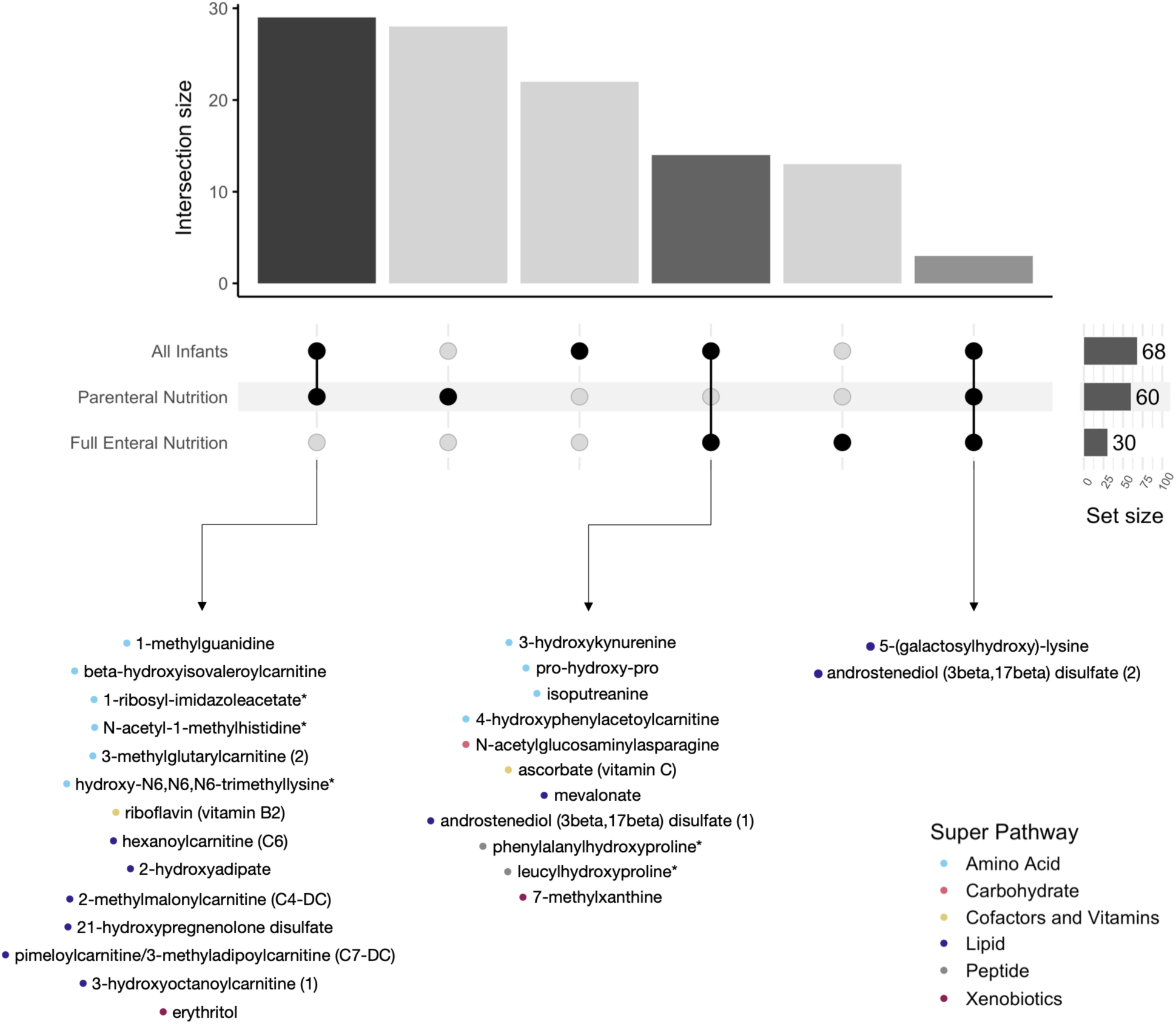
Overlap of BPD-differential metabolites across time point 2 analyses. The UpSet plot shows shared and unique BPD-differential metabolites identified in the overall time point 2 MWAS and in analyses stratified by parenteral and full enteral nutrition. Bars indicate the number of BPD-differential metabolites identified in each analysis (set size). Intersection bars show the number of metabolites shared across analyses. BPD-differential metabolites of known identity in more than one analysis are listed below their corresponding intersections. Each metabolite is accompanied by a colored dot corresponding to its super-pathway. Several BPD-differential metabolites identified among all infants at time point 2 were also identified in the nutrition-stratified analyses, including amino acid and lipid metabolites.

**Table 4.** BPD-differential metabolites of known identity unique to infants on parenteral nutrition during postnatal days 23-32 (time point 2).

| Metabolite Name | Sub-pathway | All Infants |  | Parenteral Nutrition |  | Enteral Nutrition |  |
|---|---|---|---|---|---|---|---|
|  |  | Beta (SE) | *p* | Beta (SE) | *p* | Beta (SE) | *p* |
| **ascorbic acid 3-sulfate\*** | Ascorbate and Aldarate Metabolism | -0.085 (0.046) | 0.064 | -0.268 (0.102) | 0.007 | -0.027 (0.043) | 0.465 |
| **dehydroascorbate** |  | -0.245 (0.165) | 0.106 | -0.608 (0.294) | 0.018 | -0.013 (0.255) | 0.386 |
| lactose | Disaccharides and Oligosaccharides | -0.053 (0.040) | 0.093 | -0.253 (0.123) | 0.012 | -0.020 (0.044) | 0.858 |
| adipoylcarnitine (C6-DC) | Fatty Acid Metabolism (Acyl Carnitine, Dicarboxylate) | 0.113 (0.075) | 0.865 | 0.323 (0.150) | 0.030 | 0.030 (0.063) | 0.784 |
| suberoylcarnitine (C8-DC) |  | -0.180 (0.145) | 0.492 | -0.610 (0.264) | 0.048 | 0.051 (0.190) | 0.914 |
| (S)-3-hydroxybutyrylcarnitine | Fatty Acid Metabolism (Acyl Carnitine, Hydroxy) | -0.003 (0.017) | 0.065 | -0.192 (0.089) | 0.009 | 0.006 (0.021) | 0.531 |
| 3-hydroxyhexanoylcarnitine (1) |  | -0.134 (0.072) | 0.179 | -0.397 (0.148) | 0.039 | -0.065 (0.089) | 0.645 |
| hexanoylglycine | Fatty Acid Metabolism (Acyl Glycine) | -0.053 (0.033) | 0.082 | -0.181 (0.076) | 0.039 | -0.029 (0.034) | 0.859 |
| malonylcarnitine | Fatty Acid Synthesis | -0.234 (0.214) | 0.067 | -0.616 (0.313) | 0.011 | 0.177 (0.336) | 0.947 |
| dimethylmalonic acid | Fatty Acid, Dicarboxylate | -0.065 (0.040) | 0.275 | -0.205 (0.102) | 0.049 | -0.025 (0.048) | 0.599 |
| furaneol sulfate | Food Component/Plant | -0.085 (0.049) | 0.102 | -0.211 (0.102) | 0.044 | 0.012 (0.065) | 0.607 |
| piperidine |  | -0.115 (0.069) | 0.564 | -0.259 (0.104) | 0.040 | 0.020 (0.111) | 0.612 |
| 6-oxopiperidine-2-carboxylate | Lysine Metabolism | -0.281 (0.153) | 0.216 | -0.543 (0.215) | 0.021 | 0.018 (0.267) | 0.788 |
| N6-carboxyethyllysine |  | -0.081 (0.060) | 0.567 | -0.316 (0.161) | 0.013 | -0.056 (0.070) | 0.717 |
| 1-methylhypoxanthine | Purine Metabolism, (Hypo)Xanthine/Inosine containing | 0.132 (0.107) | 0.137 | 0.327 (0.154) | 0.039 | -0.158 (0.164) | 0.958 |
| uric acid ribonucleoside* |  | -0.456 (0.240) | 0.114 | -0.619 (0.310) | 0.039 | -0.336 (0.415) | 0.886 |
| 5-methyluridine (ribothymidine) | Pyrimidine Metabolism, Uracil containing | -0.025 (0.044) | 0.131 | -0.378 (0.152) | 0.032 | 0.019 (0.053) | 0.636 |
| succinylcarnitine (C4-DC) | TCA Cycle | -0.004 (0.006) | 0.081 | -0.037 (0.018) | 0.024 | 0.004 (0.008) | 0.714 |
| **thiamin (vitamin B1)** | Thiamine Metabolism | -0.251 (0.221) | 0.065 | -0.910 (0.351) | 0.022 | 0.211 (0.331) | 0.918 |
| methylurea | Urea cycle; Arginine and Proline Metabolism | -0.107 (0.064) | 0.178 | -0.312 (0.158) | 0.050 | -0.031 (0.089) | 0.424 |
| N-monomethylarginine |  | -0.008 (0.012) | 0.214 | -0.100 (0.051) | 0.034 | -0.001 (0.014) | 0.336 |
| N,N,N-trimethyl-alanylproline betaine (TMAP) |  | -0.312 (0.179) | 0.058 | -0.568 (0.251) | 0.046 | -0.114 (0.310) | 0.418 |
| **pyridoxate** | Vitamin B6 Metabolism | -0.335 (0.181) | 0.256 | -0.528 (0.230) | 0.010 | -0.036 (0.348) | 0.524 |
| **pyridoxine (vitamin B6)** |  | -0.284 (0.180) | 0.097 | -0.453 (0.219) | 0.049 | 0.053 (0.369) | 0.730 |

*Beta* beta-coefficient; *SE* standard error; *p p*-value
Vitamins are in **boldface**.
Analysis used logistic regression for the TOLSURF and PROP cohorts separately, stratified by nutrition source (parenteral vs enteral nutrition), adjusting for gestational age, birthweight, sex, self-reported maternal race/ethnicity, and corticosteroids (>1 day), followed by a meta-analysis. A negative effect size/unit change (beta) indicates metabolites of lower abundance in BPD infants. Metabolites are grouped by sub-pathway. Sub-pathways and metabolites are listed alphabetically. Complete results are provided in the supplementary material. Corresponding *q*-values from the parenteral nutrition MWAS ranged from 0.45 to 0.73 and are provided in Supplemental Table E4.

**Table 5.** BPD-differential metabolites of known identity unique to infants on full enteral nutrition during postnatal days 23-32 (time point 2).

| Metabolite Name | Sub-pathway | All Infants |  | Parenteral Nutrition |  | Enteral Nutrition |  |
|---|---|---|---|---|---|---|---|
|  |  | Beta (SE) | *p* | Beta (SE) | *p* | Beta (SE) | *p* |
| diisopropanolamine | Chemical | 0.009 (0.090) | 0.921 | -0.105 (0.119) | 0.376 | 0.438 (0.189) | 0.021 |
| cysteinylglycine disulfide* | Glutathione Metabolism | -0.008 (0.090) | 0.931 | 0.040 (0.130) | 0.760 | -0.643 (0.291) | 0.027 |
| 2,3-dihydroxy-3-methylvalerate | Leucine, Isoleucine and Valine Metabolism | -0.006 (0.027) | 0.831 | 0.039 (0.038) | 0.304 | -0.120 (0.057) | 0.035 |
| S-methylcysteine sulfoxide | Methionine, Cysteine, SAM and Taurine Metabolism | -0.087 (0.056) | 0.119 | -0.016 (0.078) | 0.841 | -0.192 (0.083) | 0.020 |
| 4-hydroxyphenylacetate | Phenylalanine Metabolism | -0.036 (0.019) | 0.062 | -0.010 (0.037) | 0.789 | -0.053 (0.027) | 0.047 |
| 5-methylthioadenosine (MTA) | Polyamine Metabolism | 0.241 (0.125) | 0.053 | 0.085 (0.167) | 0.611 | 0.502 (0.241) | 0.037 |
| sphingosine | Sphingosines | -0.028 (0.019) | 0.146 | 0.015 (0.020) | 0.444 | -0.189 (0.096) | 0.049 |
| homocitrate | TCA Cycle | -0.054 (0.088) | 0.539 | 0.057 (0.115) | 0.621 | -0.360 (0.167) | 0.031 |
| kynurenine | Tryptophan Metabolism | -0.093 (0.088) | 0.293 | 0.064 (0.115) | 0.578 | -0.405 (0.161) | 0.012 |
| tryptophan |  | -0.197 (0.126) | 0.118 | 0.079 (0.160) | 0.622 | -0.499 (0.230) | 0.030 |

*Beta* beta-coefficient; *SE* standard error; *p p*-value
Analysis used logistic regression for the TOLSURF and PROP cohorts separately, stratified by nutrition source (parenteral vs enteral nutrition), adjusting for gestational age, birthweight, sex, self-reported maternal race/ethnicity, and corticosteroids (>1 day), followed by a meta-analysis. A negative effect size/unit change (beta) indicates metabolites of lower abundance in BPD infants. Metabolites are grouped by sub-pathway. Sub-pathways and metabolites are listed alphabetically. Complete results are provided in the supplementary material. Corresponding *q*-values from the full enteral nutrition MWAS were all 0.68 and are provided in Supplemental Table E5.

Levels of metabolites at time point 2 were asymmetric in the direction of effect in BPD, including in the analyses stratified by mode of nutrition (Figure 2B, 2C, and 2D). Among all infants, 94% of BPD-differential metabolites (64 of 68) were lower in infants with BPD at time point 2 (OR=7.1, *p*=2.2×10^-6^; Supplemental Table E2). A similar pattern was observed among infants on parenteral nutrition (57 of 60 [95%], OR=9.2, *p*=7.2×10^-7^; Supplemental Table E4) and full enteral nutrition (28 of 30 [93%], OR=6.1, *p*=0.003; Supplemental Table E5). This asymmetry was not observed at time point 1, where only 8 of 23 BPD-differential metabolites were at lower levels in infants with BPD (35%, OR=0.81, *p*=0.83). Longitudinal trends for selected BPD-differential metabolites, evaluated using a time-course design in a smaller subset of infants, showed trajectories of increasing metabolite levels after discontinuation of parenteral nutrition, with visually slower increases among infants who developed BPD (Figure 4).

See online data supplement Table E6-E10 for summary statistics on all profiled metabolites for each analysis.

### Pathway Enrichment Analysis

We performed pathway enrichment analyses for each MWAS using annotations provided by Metabolon. A total of 71 and 73 sub-pathways with ≥2 metabolites were evaluated for enrichment at time points 1 and 2, respectively, which belonged to 8 super-pathways. At time point 1, BPD-differential metabolites showed nominal enrichment of the Lactoyl Amino Acid sub-pathway (*p*=0.005, *q*=0.4, Table 6) within the Amino Acid super-pathway. At time point 2, BPD-differential metabolites were enriched in Fatty Acid Metabolism: Acyl Carnitine, Medium Chain (*p*=0.002, *q*=0.09), and Xanthine Metabolism (*p*=0.0006, *q*=0.04) (Table 6). For infants on parenteral nutrition at time point 2, BPD-differential metabolites were enriched in two lipid sub-pathways involved in fatty acid metabolism (Acyl Carnitine/Dicarboxylate [*p*=0.0002, *q*=0.007], and Acyl Carnitine/Hydroxy [*p*=0.0002, *q*=0.007]), and Vitamin B6 Metabolism (*p*=0.02, *q*=0.50) (Table 6). The Cofactors and Vitamins super-pathway was enriched for infants on parenteral nutrition at time point 2 (*p*=0.02, *q*=0.15). No other analyses showed enrichment of super-pathways (*data not shown*). For infants on full enteral nutrition at time point 2, BPD-differential metabolites were enriched for Tryptophan Metabolism (*p*=0.03, *q*=1.0) and Androgenic Steroids (*p*=0.04, *q*=1.0) (Table 6). Metabolites with fold-change nominally associated with BPD were enriched in Secondary Bile Acid Metabolism (*p*=0.008, *q*=0.59; Supplemental Table E11).

**Table 6.** Metabolomic sub-pathway enrichment analyses of BPD-differential metabolites.

| **Time Point** | **Sub-Pathway** | **Enrichment Ratio** | ***p*** | ***q*** |
|---|---|---|---|---|
| Time Point 1 | Lactoyl Amino Acid | 16.4 | 0.005 | 0.4 |
| Time Point 2 | Xanthine Metabolism | 6.3 | 0.0006 | 0.04 |
|  | Fatty acid metabolism (Acyl Carnitine, Medium Chain) | 9.1 | 0.002 | 0.09 |
| Parenteral Nutrition, Time Point 2 | Fatty Acid Metabolism (Acyl Carnitine, Dicarboxylate) | 16.6 | 0.0002 | 0.007 |
|  | Fatty Acid Metabolism (Acyl Carnitine, Hydroxy) | 16.6 | 0.0002 | 0.007 |
|  | Vitamin B6 Metabolism | 8.3 | 0.02 | 0.5 |
| Full Enteral Nutrition, Time Point 2 | Tryptophan Metabolism | 4.1 | 0.03 | 1 |
|  | Androgenic Steroids | 5.8 | 0.04 | 1 |

*p* p-value; *q* q-value representing the false discovery rate associated with each sub-pathway.
Enrichment ratio was calculated as the proportion of BPD-differential metabolites within each sub-pathway divided by the proportion of BPD-differential metabolites among all metabolites evaluated in the pathway enrichment analysis. An enrichment ratio >1 indicates overrepresentation of BPD-differential metabolites within the sub-pathway.

See online data supplement Table E11-E15 for summary statistics on all sub-pathways evaluated for each analysis.

## DISCUSSION

In this study, we characterized the urinary metabolome of 314 extremely premature infants from two cohorts within the first four weeks of life, including infants with and without a later diagnosis of BPD at 36 weeks postmenstrual age. We detected over a thousand unique metabolites representing more than 50 metabolic pathways, highlighting the breadth of physiological processes captured through performing untargeted metabolomics in a relatively non-invasive biofluid. Principal component analyses over all metabolites revealed little to no separation between infants with and without BPD, suggesting a high degree of infant variability in the urinary metabolome in this large, multi-cohort study. Levels of 86 unique individual metabolites were found to be nominally associated with BPD across time points, for which the false discovery rate should be considered when weighing their individual relevance to BPD pathophysiology. However, taken together, BPD-differential metabolites were significantly more likely to be at lower levels in the urine of infants with BPD as compared to infants without BPD, particularly during the fourth week of life and around the transition to enteral feeding. This was an unexpected finding and supports our hypothesis that infants who develop BPD are experiencing metabolic dysregulation early in life.

BPD-differential metabolites were enriched in specific metabolic pathways, including nominal enrichment of lactoyl amino acids in the second week of life, and enrichment of xanthine and fatty acid metabolism in the fourth week of life. At the earlier time point, we observed higher levels of two lactoyl amino acid metabolites in infants who developed BPD (N-lactoyl isoleucine and N-lactoyl leucine), leading to modest enrichment of the Lactoyl Amino Acid sub-pathway. N-lactoyl-amino acids are involved in a number of processes, including energy metabolism and sleep patterns [28], and higher levels have been associated with a variety of other diseases, such as gestational diabetes [29] and retinopathy of prematurity [30]. Elevated levels in infants who develop BPD could indicate greater early postnatal acidosis and a shift toward anaerobic metabolism, which may contribute to increased lung injury and poor respiratory outcomes. Previous studies have also found an association between BPD and urinary lactate levels, with lactate being a precursor of lactoyl-amino acids, including the two we identified to be at higher levels in infants with BPD. In one study, lactate was found to be higher in infants with BPD [15], whereas in another it was found to be lower in infants with BPD [16]. While we find no association between urinary lactate levels and BPD, we note that prior studies involving a smaller number of infants used a method that has lower sensitivity and is prone to degradation of lactoyl amino acids into lactate (hydrogen nuclear magnetic resonance, or H-NMR) [15,16], which may in part explain these discrepancies.

During the fourth week of life, levels of several caffeine metabolites (methylxanthines, methylurates) and acylcarnitines were lower in infants with BPD, with enrichment of the respective sub-pathways. Caffeine, an adenosine receptor antagonist commonly prescribed in the NICU to prevent apnea of prematurity, stimulates breathing, the central nervous system, and the cardiovascular system [31] and has been suggested to reduce the incidence of BPD [32]. Caffeine metabolism in preterm infants is limited by immature hepatic enzymes, while clearance is constrained by immature renal function, both of which are influenced by gestational age, post-conception age, and feeding modality [31]. Thus, lower levels of caffeine metabolites in infants who develop BPD may be attributed to these developmental limitations that reduce caffeine metabolism and renal excretion, as well as differences in dosing regimens based on body weight, caffeine blood levels, and apneic frequency. The use of caffeine was similar between infants with and without BPD, but data for dosing was not collected.

The enrichment in fatty acid metabolism was particularly evident among infants receiving parenteral nutrition as compared to infants receiving full enteral nutrition, although longitudinal trajectories suggest the role of nutrition is complex and requires further study. Acylcarnitines and carnitine are essential for energy production due to their crucial role in fatty acid oxidation, and thus they support metabolism and growth of the premature infant, which could improve respiratory outcomes. Hydroxylated fatty acids, produced by tissues and some gut microbes, may also have anti-inflammatory effects and protect against lung damage caused by inflammation, which is also a hallmark of BPD. The involvement of carnitines in BPD has similarly been observed in prior studies that used different statistical approaches, metabolomics platforms, most commonly targeted metabolomics, and that included smaller cohorts varying by gestational age, postnatal age, modes of nutrition, and biofluid [12,14,17–19], with conflicting directions of effect reported. Studies that collected samples during the first week of life or at 6 months corrected age found acylcarnitines to be higher in infants with BPD [12,14,19]. Similarly, a study evaluating metabolic differences across nutritional stages reported that the direction of carnitine dysregulation changes during these transitional periods, with higher levels predominantly in infants who developed BPD [18]. In contrast, a cross-sectional study analyzing four time points found that most acylcarnitines, particularly short- and medium-chain carnitines, were lower in infants who developed BPD at postnatal day 28, but not at postnatal days 1, 7, or 42 [17]. Our findings are consistent with those of the cross-sectional study in infants of similar postnatal age, including the replication of lower hexanoylcarnitine (C6) levels in infants who develop BPD and no differences at earlier time points. Collectively, our study, along with others, suggests that the relationship between fatty acid metabolism and BPD varies over time and by mode of nutrition (parenteral vs. enteral), a longitudinal pattern also observed for several carnitines, and that reduced fatty acid availability during the fourth week of life may be either a marker or a contributor to BPD development.

We also observed lower levels of C and B-complex vitamins during the fourth week of life in infants who developed BPD, including vitamins B_1_, B_6_, and C among infants receiving parenteral nutrition, with less pronounced differences among infants who transitioned to full enteral feeding. These findings are consistent with prior literature linking vitamin deficiencies to BPD [33–35]. Vitamin C is a cofactor in reactions regulating hypoxia-induced transcription factors and enhancing immune and anti-inflammatory responses, and a deficiency has also been attributed to cases of pulmonary hypertension [36], a common complication of BPD. B-complex vitamins support mitochondrial function [37] (vitamin B_2_), the metabolism of carbohydrates [38] (vitamins B_1_ and B_6_), amino acids [38] (vitamin B_6_), and lipids [38] (vitamin B_6_), and gut health and development [39,40]. Thus, deficiencies in B vitamins may limit energy availability in the lung, which relies on the catabolism of glucose, proteins, and lipids.

Essential vitamins are obtained through nutrition and may have varying degrees of bioavailability and concentration in parenteral vs. enteral nutrition, prompting our sub-analyses comparing infants receiving parenteral nutrition (either exclusively, transitioning to enteral feeding, or full enteral feeds for <4 days) at time point 2 vs. those receiving full enteral nutrition for at least 4 days. It is important to note that during the feeding transition, infants routinely receive both parenteral and increasing amounts of enteral nutrition, and data on daily amounts were not available. Thus, lower urinary levels of vitamin-related metabolites in infants who developed BPD could reflect impaired enteral absorption, gut immaturity, insufficient vitamin supplementation, and/or higher physiological demand in sicker preterm infants. Future studies should evaluate whether targeted nutritional supplementation, including vitamin C, B_1_, B_2_ and B_6_, may benefit infants receiving parenteral nutrition, particularly those with later or delayed transition to enteral feeds.

Vitamin deficiencies along with aberrant fatty acid metabolism could contribute to suboptimal weight gain, reduced growth, and impaired lung repair, warranting further investigation regarding their role in BPD and relationship with nutrition source. The lung and gut of infants who develop BPD are less mature developmentally for their gestational age, resulting in an increased need for respiratory, nutritional, and metabolic support. Over the first few postnatal weeks, sicker infants may have increased work of breathing and resting energy expenditure, leading to slower weight gain, less linear growth, and lower energy reserves. In addition, delays in maturational development and repair of lung structure and function could result in higher supplemental oxygen requirements, increased ventilatory pressure requirements, and transient hypoxemia with associated oxidative stress and inflammation.

Strengths of our study include the multi-cohort design, including hundreds of infants from multiple centers at two time points, thereby capturing a greater degree of individual-level variation in the urinary metabolome of extremely premature infants. A potential weakness includes the use of urine samples stored at −80°C for up to 10 years, which may have led to oxidation or degradation of certain important metabolites. In addition, our study lacked detailed information on the volume of enteral nutrition received, infant growth parameters, and caloric intake. Therefore, additional studies are needed to replicate our findings in a more contemporary cohort and to incorporate more detailed information on nutrition type.

In summary, we performed a metabolome-wide association study in extremely premature infants during the first few weeks of life and identified specific metabolites and metabolic pathways associated with the development of BPD. Our findings identify nutritional and metabolic pathways for future clinical trials of nutritional supplementation spanning lipids, essential amino acids, and vitamins B and C. Future studies are also needed to determine whether there is a causal relationship between metabolic dysregulation and the development of BPD, including the role of nutrition and nutrient absorption.

## ACKNOWLEDGEMENTS

The authors would like to acknowledge all investigators and parents of the infants for their contributions to the TOLSURF and PROP studies.

## FUNDING

This study was supported by NIH grants R01HL153429 (D.T.G., P.L.B., and C.C.), R01HL175076 (D.T.G. and R.D.H.), and R35GM158240 (R.D.H.). T.I.M.M. is a recipient of the National Science Foundation Graduate Research Fellowship. M.G. is a recipient of the Howard Hughes Medical Institute Gilliam Fellowship, Achievement Award for College Scientists Foundation Scholarship, and a UCSF Discovery Fellows Program Award. The funders had no role in the study design, data collection, analysis or interpretation, manuscript preparation, or decision to submit the manuscript for publication.

## AUTHOR CONTRIBUTIONS

Concept and design: Torgerson, Ballard, Malave-Mendez, Hernandez. Acquisition, analysis, and interpretation of data: All authors. Drafting of the manuscript: Malave-Mendez, Torgerson. Critical review of the manuscript for important intellectual content: Malave-Mendez, Ballard, Torgerson. Statistical analysis: Malave-Mendez, Torgerson, Hernandez. Obtained funding: Ballard, Torgerson, Hernandez. Administrative, technical, or material support: All authors. Supervision: Torgerson.

Dara G. Torgerson is the guarantor of the study.

## COMPETING INTERESTS

The authors declare no competing interests.

## DATA AVAILABILITY STATEMENT

Summary statistics supporting the main findings of this study are provided in the main manuscript and supplementary material. Complete summary statistics for all metabolites profiled in each MWAS will be made available after manuscript acceptance. Individual-level metabolite data are not publicly available and may be accessed upon reasonable request to Dara G. Torgerson, subject to the applicable study data access procedures and approvals.

## ETHICS APPROVAL

The research protocols for both studies were approved by the institutional review boards of the participating institutions. Written informed consent was obtained from a parent or guardian of each infant.

## PATIENT AND PUBLIC INVOLVEMENT

Patients and the public were not involved in the design, conduct, reporting, or dissemination plans of this research.

## Notes

### Competing Interest Statement

The authors have declared no competing interest.

### Clinical Trial

NCT01022580, NCT01435187

### Author Declarations

The Institutional Review Board of the University of California, San Francisco waived ethical approval for this work.

