## Supplementary Material for "Metabolome-wide Association Study of Bronchopulmonary Dysplasia in Extremely Premature Infants"

### ONLINE DATA SUPPLEMENT

***Corresponding authors:*** *Thaybeth I. Malave-Mendez, PhD, and Dara G. Torgerson, PhD. 600 16th St, University of California, San Francisco, San Francisco, CA 94158.*

| **TABLE OF CONTENTS** | **PAGES** |
|---|---|
| **Supplementary Methods** | |
| Extended Methods | 3-4 |
| References | 5 |
| **Supplementary Figures** | |
| Figure E1. Violin plots of PC1 and PC2 by BPD status. | 7 |
| **Supplementary Tables** | |
| Table E1. Summary statistics of BPD-differential metabolites at time point 1 | 8 |
| Table E2. Summary statistics of BPD-differential metabolites at time point 2 | 9 |
| Table E3. Summary statistics of metabolites with fold-change associated with BPD ($p$<0.05) | 10-11 |
| Table E4. Summary statistics of BPD-differential metabolites at time point 2 in infants on parenteral nutrition | 12 |
| Table E5. Summary statistics of BPD-differential metabolites at time point 2 in infants on enteral nutrition | 13 |
| Table E6. Summary statistics of MWAS for metabolite associations with BPD for all urinary metabolites at time point 1 (postnatal days 5-15) profiled by Metabolon | 14 |
| Table E7. Summary statistics of MWAS for metabolite associations with BPD for all urinary metabolites at time point 2 (postnatal days 23-32) profiled by Metabolon | 15 |
| Table E8. Summary statistics of MWAS for metabolite fold-change associations with BPD across all overlapping urinary metabolites profiled by Metabolon between time point 1 (postnatal days 5-15) and time point 2 (postnatal days 23-32) | 16 |
| Table E9. Summary statistics of MWAS of metabolite associations with BPD in infants on parenteral nutrition for all urinary metabolites at time point 2 (postnatal days 23-32) profiled by Metabolon | 17 |
| Table E10. Summary statistics of MWAS for metabolite associations with BPD in infants on full enteral nutrition for all urinary metabolites at time point 2 (postnatal days 23-32) profiled by Metabolon | 18 |
| Table E11. Summary statistics of sub-pathway enrichment analysis for the fold-change associations with BPD | 19-20 |
| Table E12. Summary statistics of sub-pathway enrichment analysis at time point 1 | 21-22 |
| Table E13. Summary statistics of sub-pathway enrichment analysis at time point 2 | 23-24 |
| Table E14. Summary statistics of sub-pathway enrichment analysis at time point 2 (postnatal days 23-32) for infants on parenteral nutrition | 25-26 |
| Table E15. Summary statistics of sub-pathway enrichment analysis at time point 2 (postnatal days 23-32) for infants on enteral nutrition | 27-28 |

### SUPPLEMENTARY METHODS

#### Extended Methods

**Study Description.** Infants included in our study were from the Trial of Late Surfactant for Prevention of Bronchopulmonary Dysplasia (TOLSURF, ClinicalTrials.gov, NCT01022580) and the Prematurity and Respiratory Outcomes Program (PROP, NCT01435187). Clinical data were obtained for both cohorts, including gestational age, birthweight, maternal race/ethnicity, use of corticosteroids, mode of feeding (parenteral vs enteral), and respiratory parameters, including Respiratory Severity Scores (RSS = $FiO_2$ x Mean Airway Pressure). The research protocol for both studies was approved by the institutional review boards of the participating institutions, and a parent of each infant provided written informed consent.

A total of 171 infants from TOLSURF were included in the current study, and 143 infants from PROP. Infants were selected from available samples and attempts were made to balance by bronchopulmonary dysplasia (BPD) at 36 weeks postmenstrual age, and by maternal Black, Hispanic, and non-Hispanic White race/ethnicity. TOLSURF was a double-blinded, randomized, sham-controlled trial to evaluate the effect of late surfactant on BPD and included 511 infants across 25 US centers enrolled between 2010 and 2013 [1]. Enrollment criteria included intubation and mechanical ventilation between 7 and 14 days of age and gestational age between 23-28 weeks. Infants received iNO according to the protocol followed in the Nitric Oxide (to Prevent) Chronic Lung Disease (NO CLD) trial, and BPD was assessed at 36 weeks postmenstrual age by physiological testing [2]. There was no significant difference in BPD incidence between placebo and surfactant-treated groups at 36 weeks. PROP was an observational study of 835 infants <29 weeks of gestational age from 8 US centers enrolled between 2010 and 2013 [3]. BPD was assessed at 36 weeks postmenstrual age by physiological testing using the same definition as for TOLSURF.

**Sample Collection.** Urine samples were collected from infants by placing cotton balls into the diaper for 4-8 hours and then transferred to a tube and frozen at −80°C. Samples were selected from two time points for each infant: between postnatal days 5-15 (time point 1; mean±SD 8±2) and 23-32 (time point 2; mean±SD 28±2), for a total of 628 urine samples. The total number of infants in our study was 314, including 178 with BPD and 136 without BPD (TOLSURF: N=106 with BPD and 65 without BPD; PROP: N=72 with BPD and 71 without BPD). Infants were selected based on survival at 40 weeks gestational age and availability of urine samples. In TOLSURF, infants were categorized as being on parenteral nutrition at time point 2 if they were receiving or had received parenteral nutrition within four days prior to urine sample collection, or as being on full enteral nutrition if they had not received parenteral nutrition for at least 4 days prior. In PROP, infants were categorized as being on parenteral nutrition if they had received either no or partial enteral feeds within 4 days prior to sample collection, and on enteral nutrition if they had received full enteral feeds within the same timeframe.

**Metabolomics Profiling.** Untargeted metabolomic assays were performed using Ultra High-Performance Liquid Chromatography-Tandem Mass Spectroscopy (UHPLC/MS-MS) by Metabolon Inc. as previously described [4–6]. Metabolic profiling was conducted separately for each cohort, with anchor samples included to facilitate the comparison of metabolite abundance between cohorts. Each analytic batch included a set of internal standards to adjust values for instrument variability as needed. Urine samples were normalized to osmolality, and metabolites with >30% missing values (below the limit of detection) within TOLSURF or PROP were removed from further analysis. Missing metabolite values were imputed to the minimum detectable level, and data were normalized to the median value.

**Statistical Analyses.** Principal component analysis (PCA) was performed separately at each time point to evaluate global differences in the urinary metabolome between infants with and without BPD. At time point 2, PCA was additionally performed separately among infants receiving parenteral and full enteral nutrition. A Mann-Whitney U-test was performed within each analysis to assess differences in the distributions of the first two principal components by BPD status. These analyses were performed using R version 4.5.1 (R Foundation for Statistical Computing).

A metabolome-wide association study (MWAS) was performed using multivariate logistic regression to test the association between BPD and levels of individual metabolites using the *statsmodels* package [7] in Python 3.11.4. Covariates in the model included gestational age, birth weight, sex, self-reported maternal race/ethnicity, and whether an infant was receiving corticosteroids for more than 1 day prior to urine collection. Nutrition source was included as a covariate only for time point 2. Analyses were also performed on the fold-change, calculated using the ratio of metabolite levels at time point 2 compared to time point 1. To account for study-specific and experimental batch effects, analyses were done within each cohort separately and combined using a fixed-effects meta-analysis with the *metafor* package in R

4.5.1 [8]. Sub-analyses were performed, separating infants by nutrition source at time point 2 to evaluate whether patterns of association with BPD differed by nutrition source. Metabolites with nominal $p<0.05$ were carried forward for exploratory characterization and pathway enrichment analyses and are referred to as "BPD-differential metabolites" throughout. A Fisher's exact test was used to assess whether BPD-differential metabolites were more likely than non-differential metabolites to have lower versus higher levels in infants with BPD, with odds ratios used to quantify the directional enrichment. To account for multiple testing, false discovery rates were estimated separately within each MWAS using the *qvalue* package in R [9].

Longitudinal data for selected BPD-differential metabolites from a prior study [5] were aligned to days relative to stopping parenteral nutrition and visualized using time-course plots with locally estimated scatterplot smoothing (LOESS) by BPD status. Data included untargeted metabolomics of 302 urine samples from 28 infants in TOLSURF with ≥5 time points (mean±SD 11±4) over ≥19 days (mean±SD 35±8). In this cohort, 17 (60.7%) of the infants developed BPD, and 11 (39.3%) did not develop BPD; 25 of the 28 infants were also included in the two time point study.

Pathway enrichment analysis was performed on BPD-differential metabolites for each of the MWAS comparisons performed. Metabolites of known identity from Metabolon (as of April 2025) were annotated to 8 super-pathways, including 88 sub-pathways at time point 1, and 91 sub-pathways at time point 2. Sub-pathways with fewer than 2 annotated metabolites were removed from the analysis. Pathway enrichment analysis was performed for each super-pathway and sub-pathway using a Fisher's exact test, adjusting for multiple comparisons using the *qvalue* package in R 4.5.1 [9]. The enrichment ratio was calculated as the proportion of BPD-differential metabolites within each pathway divided by the proportion of BPD-differential metabolites among all metabolites evaluated in the pathway enrichment analysis.

### SUPPLEMENTARY FIGURES

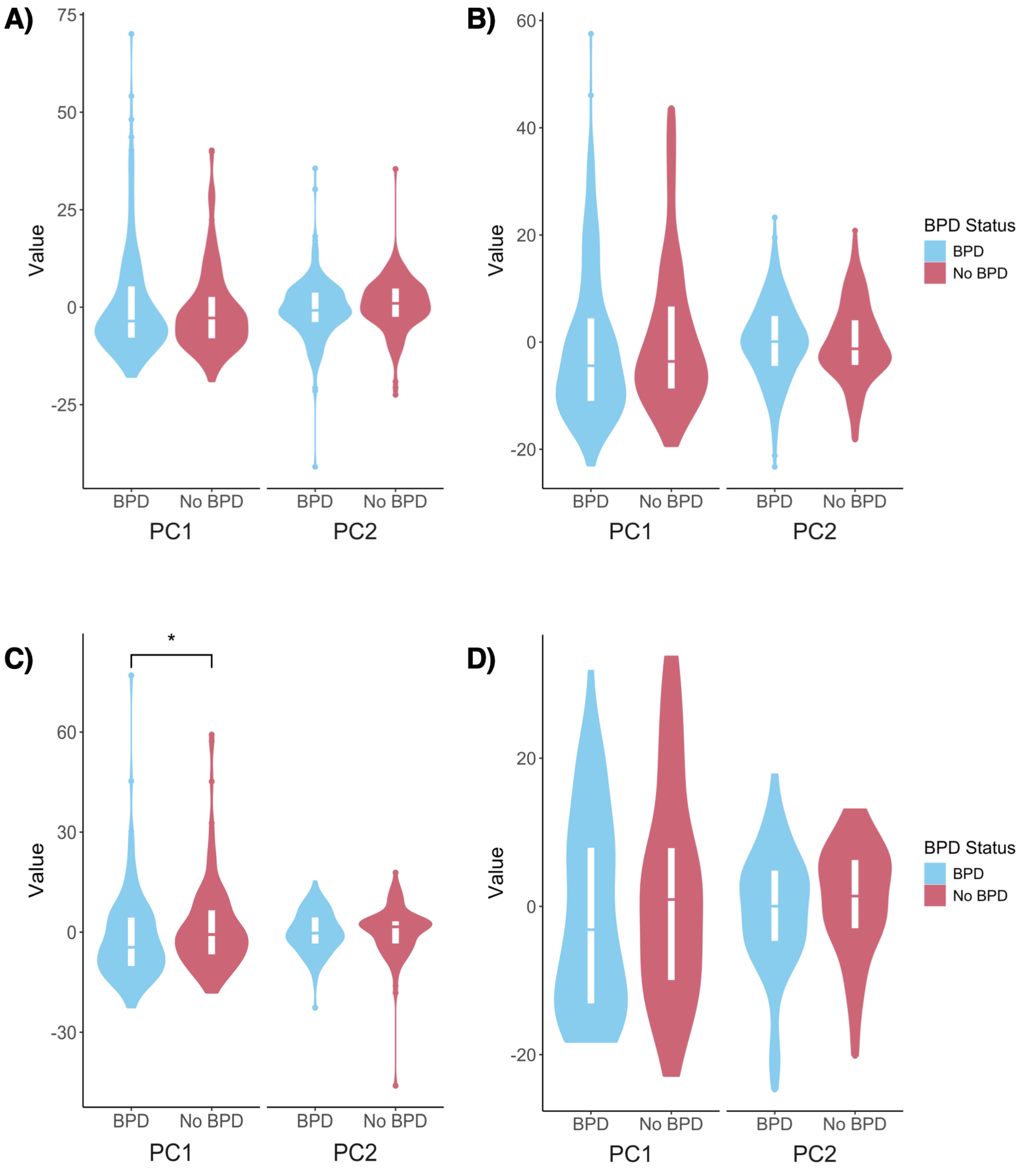


**Figure E1.** Violin plots of principal component 1 (PC1) and principal component 2 (PC2) distributions by BPD status. Comparisons are shown for: (A) all infants at time point 1 (postnatal days 5-15), (B) all infants at time point 2 (postnatal days 23-32), (C) infants on parenteral nutrition at time point 2, and (D) infants on full enteral nutrition at time point 2.

### SUPPLEMENTARY TABLES

**Table E1: Summary statistics of BPD-differential metabolites at time point 1 (postnatal days 5-15)**

| Metabolite Name | Sub-pathway | TOLSURF | | PROP | | Meta | | |
|---|---|---|---|---|---|---|---|---|
| | | Beta (SE) | *p* | Beta (SE) | *p* | Beta (SE) | *p* | *q* |
| sucrose | Disaccharides and Oligosaccharides | 0.074 (0.156) | 0.638 | -0.081 (0.030) | 0.007 | -0.076 (0.029) | 0.010 | 0.997 |
| 4-methylnonanoylcarnitine | Fatty Acid Metabolism (Acyl Carnitine, Medium Chain) | 0.021 (0.104) | 0.838 | -0.244 (0.087) | 0.005 | -0.136 (0.067) | 0.042 | 0.997 |
| methyl vanillate sulfate | Food Component/Plant | 0.137 (0.074) | 0.064 | 0.067 (0.048) | 0.155 | 0.088 (0.040) | 0.028 | 0.997 |
| menthol glucuronide | Food Component/Plant | -0.079 (0.191) | 0.680 | -0.175 (0.081) | 0.031 | -0.160 (0.075) | 0.032 | 0.997 |
| 5-aminolevulinate | Hemoglobin and Porphyrin Metabolism | 0.526 (0.326) | 0.106 | 0.756 (0.422) | 0.073 | 0.612 (0.258) | 0.018 | 0.997 |
| N-lactoyl isoleucine | Lactoyl Amino Acid | 0.389 (0.265) | 0.143 | 0.546 (0.398) | 0.171 | 0.437 (0.221) | 0.048 | 0.997 |
| N-lactoyl leucine | Lactoyl Amino Acid | 0.469 (0.290) | 0.106 | 0.458 (0.398) | 0.250 | 0.465 (0.234) | 0.047 | 0.997 |
| beta-hydroxyisovaleroylcarnitine | Leucine, Isoleucine and Valine Metabolism | -0.093 (0.248) | 0.707 | -0.512 (0.214) | 0.017 | -0.332 (0.162) | 0.040 | 0.997 |
| pipecolate | Lysine Metabolism | 0.405 (0.247) | 0.100 | 0.599 (0.377) | 0.112 | 0.463 (0.206) | 0.025 | 0.997 |
| glucuronide of C10H18O2 (11)* | Partially Characterized Molecules | 0.443 (0.216) | 0.040 | 0.276 (0.206) | 0.181 | 0.356 (0.149) | 0.017 | 0.997 |
| glucuronide of C10H18O2 (12)* | Partially Characterized Molecules | 0.284 (0.154) | 0.064 | 0.182 (0.213) | 0.394 | 0.249 (0.125) | 0.046 | 0.997 |
| guanine | Purine Metabolism, Guanine containing | 0.144 (0.161) | 0.371 | 0.904 (0.295) | 0.002 | 0.318 (0.141) | 0.024 | 0.997 |
| 3-hydroxykynurenine | Tryptophan Metabolism | 0.349 (0.175) | 0.045 | 0.388 (0.312) | 0.214 | 0.359 (0.152) | 0.019 | 0.997 |
| tyrosine | Tyrosine Metabolism | 0.163 (0.133) | 0.222 | 1.253 (0.421) | 0.003 | 0.262 (0.127) | 0.039 | 0.997 |
| caffeine | Xanthine Metabolism | -0.385 (0.205) | 0.060 | -0.226 (0.237) | 0.342 | -0.317 (0.155) | 0.041 | 0.997 |
| X-11381 | | -0.072 (0.059) | 0.218 | -0.115 (0.056) | 0.040 | -0.095 (0.041) | 0.019 | 0.997 |
| X-13431 | | -0.055 (0.143) | 0.702 | -0.540 (0.168) | 0.001 | -0.258 (0.109) | 0.018 | 0.997 |
| X-17707 | | 0.075 (0.056) | 0.185 | 0.081 (0.049) | 0.100 | 0.078 (0.037) | 0.035 | 0.997 |
| X-17763 | | 0.060 (0.057) | 0.295 | 0.080 (0.047) | 0.091 | 0.072 (0.036) | 0.049 | 0.997 |
| X-24328 | | 0.557 (0.254) | 0.028 | 0.122 (0.335) | 0.715 | 0.398 (0.202) | 0.049 | 0.997 |
| X-24359 | | 0.489 (0.196) | 0.012 | 0.252 (0.211) | 0.231 | 0.380 (0.143) | 0.008 | 0.997 |
| X-24432 | | 0.109 (0.162) | 0.502 | 0.400 (0.147) | 0.007 | 0.268 (0.109) | 0.014 | 0.997 |
| X-24801 | | -0.091 (0.184) | 0.621 | -0.436 (0.182) | 0.016 | -0.266 (0.129) | 0.040 | 0.997 |

*Meta* meta-analysis; *Beta* beta-coefficient; *SE* standard error; *p* p-value; *q* q-value

A negative beta indicates lower metabolite levels in infants with BPD.

**Table E2: Summary statistics of BPD-differential metabolites at time point 2 (postnatal days 23-32)**

Available as a separate file.

### Table E3: Summary statistics of metabolites with fold-change associated with BPD (p<0.05)

| Metabolite Name | Sub-pathway | TOLSURF | | PROP | | Meta | | |
|---|---|---|---|---|---|---|---|---|
| | | Beta (SE) | *p* | Beta (SE) | *p* | Beta (SE) | *p* | *q* |
| threonate | Ascorbate and Aldarate Metabolism | -0.002 (0.002) | 0.198 | -0.001 (0.001) | 0.106 | -0.002 (0.001) | 0.043 | 0.857 |
| hexanoylcarnitine (C6) | Fatty Acid Metabolism (Acyl Carnitine, Medium Chain) | -0.339 (0.168) | 0.043 | -0.150 (0.244) | 0.540 | -0.278 (0.138) | 0.044 | 0.857 |
| 2-hydroxystearate | Fatty Acid, Monohydroxy | -0.043 (0.039) | 0.273 | -0.092 (0.048) | 0.055 | -0.063 (0.030) | 0.039 | 0.857 |
| 2-hydroxybehenate | Fatty Acid, Monohydroxy | -0.009 (0.010) | 0.362 | -0.025 (0.010) | 0.018 | -0.017 (0.007) | 0.021 | 0.857 |
| (S)-a-amino-omega-caprolactam | Food Component/Plant | -0.264 (0.126) | 0.037 | -0.362 (0.541) | 0.504 | -0.269 (0.123) | 0.029 | 0.857 |
| carnosine | Histidine Metabolism | -0.028 (0.026) | 0.277 | -0.069 (0.030) | 0.021 | -0.046 (0.020) | 0.02 | 0.857 |
| hydantoin-5-propionate | Histidine Metabolism | -0.717 (0.710) | 0.313 | -0.209 (0.099) | 0.034 | -0.219 (0.098) | 0.025 | 0.857 |
| 2,3-dihydroxy-3-methylvalerate | Leucine, Isoleucine and Valine Metabolism | -0.012 (0.008) | 0.125 | -0.028 (0.018) | 0.121 | -0.015 (0.007) | 0.043 | 0.857 |
| glutarylcarnitine (C5-DC) | Lysine Metabolism | -0.022 (0.017) | 0.189 | -0.012 (0.008) | 0.109 | -0.014 (0.007) | 0.045 | 0.857 |
| 6-oxopiperidine-2-carboxylate | Lysine Metabolism | -0.032 (0.020) | 0.113 | -0.025 (0.020) | 0.208 | -0.029 (0.014) | 0.044 | 0.857 |
| N-acetyl-isoputreanine | Polyamine Metabolism | -0.104 (0.051) | 0.040 | -0.043 (0.038) | 0.264 | -0.065 (0.031) | 0.033 | 0.857 |
| 5alpha-pregnan-3beta,20alpha-diol disulfate | Progestin Steroids | -0.030 (0.023) | 0.187 | -0.033 (0.021) | 0.106 | -0.032 (0.015) | 0.037 | 0.857 |
| taurocholenate sulfate* | Secondary Bile Acid Metabolism | -0.055 (0.034) | 0.104 | -0.067 (0.040) | 0.095 | -0.060 (0.026) | 0.02 | 0.857 |
| glycohyocholate | Secondary Bile Acid Metabolism | -0.217 (0.088) | 0.014 | -0.050 (0.059) | 0.398 | -0.102 (0.049) | 0.039 | 0.857 |
| tricarballylate | TCA Cycle | -0.133 (0.088) | 0.131 | -0.169 (0.130) | 0.195 | -0.145 (0.073) | 0.048 | 0.857 |
| gamma-CEHC sulfate | Tocopherol Metabolism | -0.227 (0.140) | 0.105 | -0.166 (0.122) | 0.171 | -0.192 (0.092) | 0.036 | 0.857 |
| 3-indoxyl sulfate | Tryptophan Metabolism | -0.036 (0.022) | 0.103 | -0.026 (0.020) | 0.193 | -0.031 (0.015) | 0.039 | 0.857 |
| tryptophan betaine | Tryptophan Metabolism | -0.089 (0.064) | 0.165 | -0.090 (0.043) | 0.036 | -0.090 (0.036) | 0.012 | 0.857 |
| N-methylproline | Urea cycle; Arginine and Proline Metabolism | 0.073 (0.057) | 0.196 | 0.036 (0.020) | 0.074 | 0.040 (0.019) | 0.034 | 0.857 |
| X-11612 | | -0.071 (0.037) | 0.056 | -0.099 (0.071) | 0.16 | -0.077 (0.033) | 0.019 | 0.857 |
| X-12117 | | -0.236 (0.139) | 0.089 | -0.344 (0.294) | 0.242 | -0.256 (0.126) | 0.042 | 0.857 |
| X-12844 | | -0.013 (0.021) | 0.535 | -0.017 (0.008) | 0.044 | -0.016 (0.008) | 0.035 | 0.857 |

| | | | | | | | |
|---|---|---|---|---|---|---|---|
| X-15486 | -0.050 (0.029) | 0.091 | -0.024 (0.017) | 0.152 | -0.030 (0.014) | 0.038 | 0.857 |
| X-17357 | -0.034 (0.040) | 0.391 | -0.055 (0.029) | 0.059 | -0.048 (0.024) | 0.043 | 0.857 |
| X-17674 | -0.055 (0.036) | 0.127 | -0.215 (0.105) | 0.041 | -0.072 (0.034) | 0.035 | 0.857 |
| X-21815 | -0.023 (0.011) | 0.033 | -0.009 (0.009) | 0.345 | -0.015 (0.007) | 0.036 | 0.857 |
| X-24669 | -0.015 (0.009) | 0.084 | -0.012 (0.010) | 0.239 | -0.014 (0.007) | 0.038 | 0.857 |
| X-25422 | -0.154 (0.075) | 0.039 | -0.804 (0.319) | 0.012 | -0.188 (0.073) | 0.01 | 0.857 |

*Meta* meta-analysis; *Beta* beta-coefficient; *SE* standard error; *p* p-value; *q* q-value

A negative beta indicates a lower fold-change from time point 1 to time point 2 in infants with BPD.

**Table E4: Summary statistics of BPD-differential metabolites at time point 2 (postnatal days 23-32) in infants on parenteral nutrition**

Available as a separate file.

**Table E5: Summary statistics of BPD-differential metabolites at time point 2 (postnatal days 23-32) in infants on full enteral nutrition**

Available as a separate file

**Table E6: Summary Statistics of MWAS for Metabolite Associations with BPD for all Urinary Metabolites at Time Point 1 (postnatal days 5-15) Profiled by Metabolon**

Available as a separate file.

**Table E7: Summary Statistics of MWAS for Metabolite Associations with BPD for all Urinary Metabolites at Time Point 2 (postnatal days 23-32) Profiled by Metabolon**

Available as a separate file.

**Table E8: Summary statistics of MWAS for metabolite fold-change associations with BPD across all overlapping urinary metabolites profiled by Metabolon between time point 1 (postnatal days 5-15) and time point 2 (postnatal days 23-32)**

Available as a separate file.

**Table E9: Summary Statistics of MWAS for Metabolite Associations with BPD for all Urinary Metabolites at Time Point 2 (postnatal days 23-32) Profiled by Metabolon in Infants on Parenteral Nutrition**

Available as a separate file.

**Table E10: Summary Statistics of MWAS for Metabolite Associations with BPD for all Urinary Metabolites at Time Point 2 (postnatal days 23-32) Profiled by Metabolon in Infants on Full Enteral Nutrition**

Available as a separate file.

**Table E11: Summary statistics of sub-pathway enrichment analysis for the fold-change associations with BPD**

| Sub-Pathway | Super-Pathway | Odds Ratio | Enrichment Ratio | *p* | *q* |
|---|---|---|---|---|---|
| Secondary Bile Acid Metabolism | Lipid | 23.608 | 13.137 | 0.008 | 0.591 |
| Fatty Acid, Monohydroxy | Lipid | 7.000 | 5.474 | 0.048 | 1.000 |
| Progestin Steroids | Lipid | 33.556 | 16.421 | 0.060 | 1.000 |
| Fatty Acid Metabolism (Acyl Carnitine, Medium Chain) | Lipid | 11.148 | 8.211 | 0.117 | 1.000 |
| Tryptophan Metabolism | Amino Acid | 3.837 | 3.284 | 0.120 | 1.000 |
| Histidine Metabolism | Amino Acid | 3.441 | 2.986 | 0.141 | 1.000 |
| Tocopherol Metabolism | Cofactors and Vitamins | 8.347 | 6.568 | 0.144 | 1.000 |
| Lysine Metabolism | Amino Acid | 3.272 | 2.856 | 0.152 | 1.000 |
| Ascorbate and Aldarate Metabolism | Cofactors and Vitamins | 4.746 | 4.105 | 0.220 | 1.000 |
| Polyamine Metabolism | Amino Acid | 3.000 | 2.737 | 0.312 | 1.000 |
| TCA Cycle | Energy | 2.530 | 2.346 | 0.354 | 1.000 |
| Urea cycle; Arginine and Proline Metabolism | Amino Acid | 1.289 | 1.263 | 0.560 | 1.000 |
| Leucine, Isoleucine and Valine Metabolism | Amino Acid | 1.103 | 1.095 | 0.613 | 1.000 |
| Acetylated Peptides | Peptide | 0.000 | 0.000 | 1.000 | 1.000 |
| Advanced Glycation End-product | Carbohydrate | 0.000 | 0.000 | 1.000 | 1.000 |
| Alanine and Aspartate Metabolism | Amino Acid | 0.000 | 0.000 | 1.000 | 1.000 |
| Aminosugar Metabolism | Carbohydrate | 0.000 | 0.000 | 1.000 | 1.000 |
| Androgenic Steroids | Lipid | 0.000 | 0.000 | 1.000 | 1.000 |
| Bacterial/Fungal | Xenobiotics | 0.000 | 0.000 | 1.000 | 1.000 |
| Benzoate Metabolism | Xenobiotics | 0.000 | 0.000 | 1.000 | 1.000 |
| Carnitine Metabolism | Lipid | 0.000 | 0.000 | 1.000 | 1.000 |
| Chemical | Xenobiotics | 0.000 | 0.000 | 1.000 | 1.000 |
| Corticosteroids | Lipid | 0.000 | 0.000 | 1.000 | 1.000 |
| Creatine Metabolism | Amino Acid | 0.000 | 0.000 | 1.000 | 1.000 |
| Dipeptide | Peptide | 0.000 | 0.000 | 1.000 | 1.000 |
| Dipeptide Derivative | Peptide | 0.000 | 0.000 | 1.000 | 1.000 |
| Disaccharides and Oligosaccharides | Carbohydrate | 0.000 | 0.000 | 1.000 | 1.000 |
| Drug - Analgesics, Anesthetics | Xenobiotics | 0.000 | 0.000 | 1.000 | 1.000 |
| Drug - Topical Agents | Xenobiotics | 0.000 | 0.000 | 1.000 | 1.000 |
| Fatty Acid Metabolism (Acyl Carnitine, Dicarboxylate) | Lipid | 0.000 | 0.000 | 1.000 | 1.000 |
| Fatty Acid Metabolism (Acyl Carnitine, Hydroxy) | Lipid | 0.000 | 0.000 | 1.000 | 1.000 |
| Fatty Acid Metabolism (Acyl Glutamine) | Lipid | 0.000 | 0.000 | 1.000 | 1.000 |
| Fatty Acid Metabolism (Acyl Glycine) | Lipid | 0.000 | 0.000 | 1.000 | 1.000 |
| Fatty Acid Metabolism (also BCAA Metabolism) | Lipid | 0.000 | 0.000 | 1.000 | 1.000 |
| Fatty Acid Synthesis | Lipid | 0.000 | 0.000 | 1.000 | 1.000 |
| Fatty Acid, Branched | Lipid | 0.000 | 0.000 | 1.000 | 1.000 |
| Fatty Acid, Dicarboxylate | Lipid | 0.000 | 0.000 | 1.000 | 1.000 |

| | | | | | |
|---|---|---|---|---|---|
| Fatty Acid, Dihydroxy | Lipid | 0.000 | 0.000 | 1.000 | 1.000 |
| Food Component/Plant | Xenobiotics | 0.995 | 0.995 | 1.000 | 1.000 |
| Fructose, Mannose and Galactose Metabolism | Carbohydrate | 0.000 | 0.000 | 1.000 | 1.000 |
| Gamma-glutamyl Amino Acid | Peptide | 0.000 | 0.000 | 1.000 | 1.000 |
| Glutamate Metabolism | Amino Acid | 0.000 | 0.000 | 1.000 | 1.000 |
| Glutathione Metabolism | Amino Acid | 0.000 | 0.000 | 1.000 | 1.000 |
| Glycerolipid Metabolism | Lipid | 0.000 | 0.000 | 1.000 | 1.000 |
| Glycine, Serine and Threonine Metabolism | Amino Acid | 0.000 | 0.000 | 1.000 | 1.000 |
| Glycolysis, Gluconeogenesis, and Pyruvate Metabolism | Carbohydrate | 0.000 | 0.000 | 1.000 | 1.000 |
| Guanidino and Acetamido Metabolism | Amino Acid | 0.000 | 0.000 | 1.000 | 1.000 |
| Lactoyl Amino Acid | Amino Acid | 0.000 | 0.000 | 1.000 | 1.000 |
| Long Chain Monounsaturated Fatty Acid | Lipid | 0.000 | 0.000 | 1.000 | 1.000 |
| Long Chain Polyunsaturated Fatty Acid (n3 and n6) | Lipid | 0.000 | 0.000 | 1.000 | 1.000 |
| Long Chain Saturated Fatty Acid | Lipid | 0.000 | 0.000 | 1.000 | 1.000 |
| Methionine, Cysteine, SAM and Taurine Metabolism | Amino Acid | 0.000 | 0.000 | 1.000 | 1.000 |
| Mevalonate Metabolism | Lipid | 0.000 | 0.000 | 1.000 | 1.000 |
| Nicotinate and Nicotinamide Metabolism | Cofactors and Vitamins | 0.000 | 0.000 | 1.000 | 1.000 |
| Pantothenate and CoA Metabolism | Cofactors and Vitamins | 0.000 | 0.000 | 1.000 | 1.000 |
| Pentose Metabolism | Carbohydrate | 0.000 | 0.000 | 1.000 | 1.000 |
| Phenylalanine Metabolism | Amino Acid | 0.000 | 0.000 | 1.000 | 1.000 |
| Phospholipid Metabolism | Lipid | 0.000 | 0.000 | 1.000 | 1.000 |
| Pregnenolone Steroids | Lipid | 0.000 | 0.000 | 1.000 | 1.000 |
| Primary Bile Acid Metabolism | Lipid | 0.000 | 0.000 | 1.000 | 1.000 |
| Pterin Metabolism | Cofactors and Vitamins | 0.000 | 0.000 | 1.000 | 1.000 |
| Purine Metabolism, (Hypo)Xanthine/Inosine containing | Nucleotide | 0.000 | 0.000 | 1.000 | 1.000 |
| Purine Metabolism, Adenine containing | Nucleotide | 0.000 | 0.000 | 1.000 | 1.000 |
| Purine Metabolism, Guanine containing | Nucleotide | 0.000 | 0.000 | 1.000 | 1.000 |
| Pyrimidine Metabolism, Cytidine containing | Nucleotide | 0.000 | 0.000 | 1.000 | 1.000 |
| Pyrimidine Metabolism, Orotate containing | Nucleotide | 0.000 | 0.000 | 1.000 | 1.000 |
| Pyrimidine Metabolism, Thymine containing | Nucleotide | 0.000 | 0.000 | 1.000 | 1.000 |
| Pyrimidine Metabolism, Uracil containing | Nucleotide | 0.000 | 0.000 | 1.000 | 1.000 |
| Tyrosine Metabolism | Amino Acid | 0.000 | 0.000 | 1.000 | 1.000 |
| Vitamin B6 Metabolism | Cofactors and Vitamins | 0.000 | 0.000 | 1.000 | 1.000 |
| Xanthine Metabolism | Xenobiotics | 0.000 | 0.000 | 1.000 | 1.000 |

*p* p-value; *q* q-value

An enrichment ratio >1 indicates overrepresentation of BPD-differential metabolites within the sub-pathway.

**Table E12: Summary statistics of sub-pathway enrichment analysis at time point 1 (postnatal days 5-15)**

| Sub-Pathway | Super-Pathway | Odds Ratio | Enrichment Ratio | *p* | *q* |
|---|---|---|---|---|---|
| Lactoyl Amino Acid | Amino Acid | 28.318 | 16.410 | 0.005 | 0.388 |
| Disaccharides and Oligosaccharides | Carbohydrate | 17.333 | 12.308 | 0.079 | 1.000 |
| Fatty Acid Metabolism (Acyl Carnitine, Medium Chain) | Lipid | 17.333 | 12.308 | 0.079 | 1.000 |
| Xanthine Metabolism | Xenobiotics | 8.625 | 7.033 | 0.134 | 1.000 |
| Purine Metabolism, Guanine containing | Nucleotide | 7.381 | 6.154 | 0.152 | 1.000 |
| Food Component/Plant | Xenobiotics | 3.273 | 2.813 | 0.155 | 1.000 |
| Tryptophan Metabolism | Amino Acid | 2.667 | 2.462 | 0.341 | 1.000 |
| Lysine Metabolism | Amino Acid | 2.292 | 2.140 | 0.381 | 1.000 |
| Tyrosine Metabolism | Amino Acid | 1.718 | 1.641 | 0.468 | 1.000 |
| Leucine, Isoleucine and Valine Metabolism | Amino Acid | 1.602 | 1.538 | 0.490 | 1.000 |
| Acetylated Peptides | Peptide | 0.000 | 0.000 | 1.000 | 1.000 |
| Advanced Glycation End-product | Carbohydrate | 0.000 | 0.000 | 1.000 | 1.000 |
| Alanine and Aspartate Metabolism | Amino Acid | 0.000 | 0.000 | 1.000 | 1.000 |
| Aminosugar Metabolism | Carbohydrate | 0.000 | 0.000 | 1.000 | 1.000 |
| Androgenic Steroids | Lipid | 0.000 | 0.000 | 1.000 | 1.000 |
| Ascorbate and Aldarate Metabolism | Cofactors and Vitamins | 0.000 | 0.000 | 1.000 | 1.000 |
| Bacterial/Fungal | Xenobiotics | 0.000 | 0.000 | 1.000 | 1.000 |
| Benzoate Metabolism | Xenobiotics | 0.000 | 0.000 | 1.000 | 1.000 |
| Carnitine Metabolism | Lipid | 0.000 | 0.000 | 1.000 | 1.000 |
| Chemical | Xenobiotics | 0.000 | 0.000 | 1.000 | 1.000 |
| Corticosteroids | Lipid | 0.000 | 0.000 | 1.000 | 1.000 |
| Creatine Metabolism | Amino Acid | 0.000 | 0.000 | 1.000 | 1.000 |
| Dipeptide | Peptide | 0.000 | 0.000 | 1.000 | 1.000 |
| Dipeptide Derivative | Peptide | 0.000 | 0.000 | 1.000 | 1.000 |
| Drug - Analgesics, Anesthetics | Xenobiotics | 0.000 | 0.000 | 1.000 | 1.000 |
| Drug - Topical Agents | Xenobiotics | 0.000 | 0.000 | 1.000 | 1.000 |
| Fatty Acid Metabolism (Acyl Carnitine, Dicarboxylate) | Lipid | 0.000 | 0.000 | 1.000 | 1.000 |
| Fatty Acid Metabolism (Acyl Carnitine, Hydroxy) | Lipid | 0.000 | 0.000 | 1.000 | 1.000 |
| Fatty Acid Metabolism (Acyl Glutamine) | Lipid | 0.000 | 0.000 | 1.000 | 1.000 |
| Fatty Acid Metabolism (Acyl Glycine) | Lipid | 0.000 | 0.000 | 1.000 | 1.000 |
| Fatty Acid Metabolism (also BCAA Metabolism) | Lipid | 0.000 | 0.000 | 1.000 | 1.000 |
| Fatty Acid Synthesis | Lipid | 0.000 | 0.000 | 1.000 | 1.000 |
| Fatty Acid, Branched | Lipid | 0.000 | 0.000 | 1.000 | 1.000 |
| Fatty Acid, Dicarboxylate | Lipid | 0.000 | 0.000 | 1.000 | 1.000 |
| Fatty Acid, Dihydroxy | Lipid | 0.000 | 0.000 | 1.000 | 1.000 |
| Fatty Acid, Monohydroxy | Lipid | 0.000 | 0.000 | 1.000 | 1.000 |
| Fructose, Mannose and Galactose Metabolism | Carbohydrate | 0.000 | 0.000 | 1.000 | 1.000 |

| | | | | | |
|---|---|---|---|---|---|
| Gamma-glutamyl Amino Acid | Peptide | 0.000 | 0.000 | 1.000 | 1.000 |
| Glutamate Metabolism | Amino Acid | 0.000 | 0.000 | 1.000 | 1.000 |
| Glutathione Metabolism | Amino Acid | 0.000 | 0.000 | 1.000 | 1.000 |
| Glycerolipid Metabolism | Lipid | 0.000 | 0.000 | 1.000 | 1.000 |
| Glycine, Serine and Threonine Metabolism | Amino Acid | 0.000 | 0.000 | 1.000 | 1.000 |
| Glycolysis, Gluconeogenesis, and Pyruvate Metabolism | Carbohydrate | 0.000 | 0.000 | 1.000 | 1.000 |
| Guanidino and Acetamido Metabolism | Amino Acid | 0.000 | 0.000 | 1.000 | 1.000 |
| Histidine Metabolism | Amino Acid | 0.000 | 0.000 | 1.000 | 1.000 |
| Long Chain Monounsaturated Fatty Acid | Lipid | 0.000 | 0.000 | 1.000 | 1.000 |
| Long Chain Polyunsaturated Fatty Acid (n3 and n6) | Lipid | 0.000 | 0.000 | 1.000 | 1.000 |
| Long Chain Saturated Fatty Acid | Lipid | 0.000 | 0.000 | 1.000 | 1.000 |
| Methionine, Cysteine, SAM and Taurine Metabolism | Amino Acid | 0.000 | 0.000 | 1.000 | 1.000 |
| Mevalonate Metabolism | Lipid | 0.000 | 0.000 | 1.000 | 1.000 |
| Nicotinate and Nicotinamide Metabolism | Cofactors and Vitamins | 0.000 | 0.000 | 1.000 | 1.000 |
| Pantothenate and CoA Metabolism | Cofactors and Vitamins | 0.000 | 0.000 | 1.000 | 1.000 |
| Pentose Metabolism | Carbohydrate | 0.000 | 0.000 | 1.000 | 1.000 |
| Phenylalanine Metabolism | Amino Acid | 0.000 | 0.000 | 1.000 | 1.000 |
| Phospholipid Metabolism | Lipid | 0.000 | 0.000 | 1.000 | 1.000 |
| Polyamine Metabolism | Amino Acid | 0.000 | 0.000 | 1.000 | 1.000 |
| Pregnenolone Steroids | Lipid | 0.000 | 0.000 | 1.000 | 1.000 |
| Primary Bile Acid Metabolism | Lipid | 0.000 | 0.000 | 1.000 | 1.000 |
| Progestin Steroids | Lipid | 0.000 | 0.000 | 1.000 | 1.000 |
| Pterin Metabolism | Cofactors and Vitamins | 0.000 | 0.000 | 1.000 | 1.000 |
| Purine Metabolism, (Hypo)Xanthine/Inosine containing | Nucleotide | 0.000 | 0.000 | 1.000 | 1.000 |
| Purine Metabolism, Adenine containing | Nucleotide | 0.000 | 0.000 | 1.000 | 1.000 |
| Pyrimidine Metabolism, Cytidine containing | Nucleotide | 0.000 | 0.000 | 1.000 | 1.000 |
| Pyrimidine Metabolism, Orotate containing | Nucleotide | 0.000 | 0.000 | 1.000 | 1.000 |
| Pyrimidine Metabolism, Thymine containing | Nucleotide | 0.000 | 0.000 | 1.000 | 1.000 |
| Pyrimidine Metabolism, Uracil containing | Nucleotide | 0.000 | 0.000 | 1.000 | 1.000 |
| Secondary Bile Acid Metabolism | Lipid | 0.000 | 0.000 | 1.000 | 1.000 |
| TCA Cycle | Energy | 0.000 | 0.000 | 1.000 | 1.000 |
| Tocopherol Metabolism | Cofactors and Vitamins | 0.000 | 0.000 | 1.000 | 1.000 |
| Urea cycle; Arginine and Proline Metabolism | Amino Acid | 0.000 | 0.000 | 1.000 | 1.000 |
| Vitamin B6 Metabolism | Cofactors and Vitamins | 0.000 | 0.000 | 1.000 | 1.000 |

*p* p-value; *q* q-value

181 An enrichment ratio >1 indicates overrepresentation of BPD-differential metabolites within the sub-pathway.

**Table E13: Summary statistics of sub-pathway enrichment analysis at time point 2 (postnatal days 23-32)**

| Sub-Pathway | Super-Pathway | Odds Ratio | Enrichment Ratio | *p* | q |
|---|---|---|---|---|---|
| Xanthine Metabolism | Xenobiotics | 11.227 | 6.288 | 0.001 | 0.042 |
| Fatty Acid Metabolism (Acyl Carnitine, Medium Chain) | Lipid | 22.610 | 9.055 | 0.002 | 0.090 |
| Bacterial/Fungal | Xenobiotics | 14.395 | 7.545 | 0.128 | 1.000 |
| Dipeptide Derivative | Peptide | 14.395 | 7.545 | 0.128 | 1.000 |
| Leucine, Isoleucine and Valine Metabolism | Amino Acid | 2.285 | 2.012 | 0.130 | 1.000 |
| Androgenic Steroids | Lipid | 3.643 | 3.018 | 0.138 | 1.000 |
| Fatty Acid Metabolism (Acyl Carnitine, Dicarboxylate) | Lipid | 7.186 | 5.030 | 0.186 | 1.000 |
| Fatty Acid Metabolism (Acyl Carnitine, Hydroxy) | Lipid | 7.186 | 5.030 | 0.186 | 1.000 |
| Fatty Acid Metabolism (also BCAA Metabolism) | Lipid | 7.186 | 5.030 | 0.186 | 1.000 |
| Guanidino and Acetamido Metabolism | Amino Acid | 7.186 | 5.030 | 0.186 | 1.000 |
| Mevalonate Metabolism | Lipid | 7.186 | 5.030 | 0.186 | 1.000 |
| Histidine Metabolism | Amino Acid | 2.195 | 1.968 | 0.190 | 1.000 |
| Lysine Metabolism | Amino Acid | 2.087 | 1.886 | 0.208 | 1.000 |
| Pregnenolone Steroids | Lipid | 2.380 | 2.156 | 0.383 | 1.000 |
| Benzoate Metabolism | Xenobiotics | 0.000 | 0.000 | 0.389 | 1.000 |
| Aminosugar Metabolism | Carbohydrate | 2.037 | 1.886 | 0.424 | 1.000 |
| Ascorbate and Aldarate Metabolism | Cofactors and Vitamins | 2.037 | 1.886 | 0.424 | 1.000 |
| Dipeptide | Peptide | 1.779 | 1.677 | 0.463 | 1.000 |
| Polyamine Metabolism | Amino Acid | 1.288 | 1.258 | 0.564 | 1.000 |
| TCA Cycle | Energy | 0.000 | 0.000 | 0.615 | 1.000 |
| Tryptophan Metabolism | Amino Acid | 1.506 | 1.437 | 0.643 | 1.000 |
| Chemical | Xenobiotics | 1.429 | 1.372 | 0.651 | 1.000 |
| Tyrosine Metabolism | Amino Acid | 0.474 | 0.503 | 0.713 | 1.000 |
| Food Component/Plant | Xenobiotics | 0.401 | 0.431 | 0.722 | 1.000 |
| Acetylated Peptides | Peptide | 0.000 | 0.000 | 1.000 | 1.000 |
| Advanced Glycation End-product | Carbohydrate | 0.000 | 0.000 | 1.000 | 1.000 |
| Alanine and Aspartate Metabolism | Amino Acid | 0.000 | 0.000 | 1.000 | 1.000 |
| Carnitine Metabolism | Lipid | 0.000 | 0.000 | 1.000 | 1.000 |
| Corticosteroids | Lipid | 0.000 | 0.000 | 1.000 | 1.000 |
| Creatine Metabolism | Amino Acid | 0.000 | 0.000 | 1.000 | 1.000 |
| Disaccharides and Oligosaccharides | Carbohydrate | 0.000 | 0.000 | 1.000 | 1.000 |
| Drug - Analgesics, Anesthetics | Xenobiotics | 0.000 | 0.000 | 1.000 | 1.000 |
| Drug - Topical Agents | Xenobiotics | 0.000 | 0.000 | 1.000 | 1.000 |
| Fatty Acid Metabolism (Acyl Glutamine) | Lipid | 0.000 | 0.000 | 1.000 | 1.000 |
| Fatty Acid Metabolism (Acyl Glycine) | Lipid | 0.000 | 0.000 | 1.000 | 1.000 |
| Fatty Acid Synthesis | Lipid | 0.000 | 0.000 | 1.000 | 1.000 |
| Fatty Acid, Branched | Lipid | 0.000 | 0.000 | 1.000 | 1.000 |

| | | | | | |
|---|---|---|---|---|---|
| Fatty Acid, Dicarboxylate | Lipid | 0.698 | 0.719 | 1.000 | 1.000 |
| Fatty Acid, Dihydroxy | Lipid | 0.000 | 0.000 | 1.000 | 1.000 |
| Fatty Acid, Monohydroxy | Lipid | 0.000 | 0.000 | 1.000 | 1.000 |
| Fructose, Mannose and Galactose Metabolism | Carbohydrate | 0.000 | 0.000 | 1.000 | 1.000 |
| Gamma-glutamyl Amino Acid | Peptide | 0.000 | 0.000 | 1.000 | 1.000 |
| Glutamate Metabolism | Amino Acid | 0.000 | 0.000 | 1.000 | 1.000 |
| Glutathione Metabolism | Amino Acid | 0.000 | 0.000 | 1.000 | 1.000 |
| Glycerolipid Metabolism | Lipid | 0.000 | 0.000 | 1.000 | 1.000 |
| Glycine, Serine and Threonine Metabolism | Amino Acid | 0.000 | 0.000 | 1.000 | 1.000 |
| Glycolysis, Gluconeogenesis, and Pyruvate Metabolism | Carbohydrate | 0.000 | 0.000 | 1.000 | 1.000 |
| Lactoyl Amino Acid | Amino Acid | 0.000 | 0.000 | 1.000 | 1.000 |
| Long Chain Monounsaturated Fatty Acid | Lipid | 0.000 | 0.000 | 1.000 | 1.000 |
| Long Chain Polyunsaturated Fatty Acid (n3 and n6) | Lipid | 0.000 | 0.000 | 1.000 | 1.000 |
| Long Chain Saturated Fatty Acid | Lipid | 0.000 | 0.000 | 1.000 | 1.000 |
| Methionine, Cysteine, SAM and Taurine Metabolism | Amino Acid | 0.663 | 0.686 | 1.000 | 1.000 |
| Nicotinate and Nicotinamide Metabolism | Cofactors and Vitamins | 0.000 | 0.000 | 1.000 | 1.000 |
| Pantothenate and CoA Metabolism | Cofactors and Vitamins | 0.000 | 0.000 | 1.000 | 1.000 |
| Pentose Metabolism | Carbohydrate | 0.000 | 0.000 | 1.000 | 1.000 |
| Phenylalanine Metabolism | Amino Acid | 0.000 | 0.000 | 1.000 | 1.000 |
| Phospholipid Metabolism | Lipid | 0.000 | 0.000 | 1.000 | 1.000 |
| Primary Bile Acid Metabolism | Lipid | 0.000 | 0.000 | 1.000 | 1.000 |
| Progestin Steroids | Lipid | 0.000 | 0.000 | 1.000 | 1.000 |
| Pterin Metabolism | Cofactors and Vitamins | 0.000 | 0.000 | 1.000 | 1.000 |
| Purine Metabolism, (Hypo)Xanthine/Inosine containing | Nucleotide | 0.000 | 0.000 | 1.000 | 1.000 |
| Purine Metabolism, Adenine containing | Nucleotide | 0.000 | 0.000 | 1.000 | 1.000 |
| Purine Metabolism, Guanine containing | Nucleotide | 0.000 | 0.000 | 1.000 | 1.000 |
| Pyrimidine Metabolism, Cytidine containing | Nucleotide | 0.000 | 0.000 | 1.000 | 1.000 |
| Pyrimidine Metabolism, Orotate containing | Nucleotide | 0.000 | 0.000 | 1.000 | 1.000 |
| Pyrimidine Metabolism, Thymine containing | Nucleotide | 0.000 | 0.000 | 1.000 | 1.000 |
| Pyrimidine Metabolism, Uracil containing | Nucleotide | 1.007 | 1.006 | 1.000 | 1.000 |
| Secondary Bile Acid Metabolism | Lipid | 0.000 | 0.000 | 1.000 | 1.000 |
| Tetrahydrobiopterin Metabolism | Cofactors and Vitamins | 0.000 | 0.000 | 1.000 | 1.000 |
| Thiamine Metabolism | Cofactors and Vitamins | 0.000 | 0.000 | 1.000 | 1.000 |
| Tocopherol Metabolism | Cofactors and Vitamins | 0.000 | 0.000 | 1.000 | 1.000 |
| Urea cycle; Arginine and Proline Metabolism | Amino Acid | 0.553 | 0.580 | 1.000 | 1.000 |
| Vitamin B6 Metabolism | Cofactors and Vitamins | 0.000 | 0.000 | 1.000 | 1.000 |

*p* p-value; *q* q-value

182 An enrichment ratio >1 indicates overrepresentation of BPD-differential metabolites within the sub-pathway.

**Table E14: Summary statistics of sub-pathway enrichment analysis at time point 2 (postnatal days 23-32) for infants on parenteral nutrition**

| Sub-Pathway | Super-Pathway | Odds Ratio | Enrichment Ratio | *p* | *q* |
|---|---|---|---|---|---|
| Fatty Acid Metabolism (Acyl Carnitine, Dicarboxylate) | Lipid | Infinite | 16.600 | 0.0002 | 0.007 |
| Fatty Acid Metabolism (Acyl Carnitine, Hydroxy) | Lipid | Infinite | 16.600 | 0.0002 | 0.007 |
| Vitamin B6 Metabolism | Cofactors and Vitamins | 16.368 | 8.300 | 0.020 | 0.479 |
| Lysine Metabolism | Amino Acid | 3.356 | 2.767 | 0.050 | 0.911 |
| Ascorbate and Aldarate Metabolism | Cofactors and Vitamins | 5.421 | 4.150 | 0.079 | 1.000 |
| Purine Metabolism, (Hypo)Xanthine/Inosine containing | Nucleotide | 4.639 | 3.689 | 0.097 | 1.000 |
| Fatty Acid Synthesis | Lipid | 15.974 | 8.300 | 0.117 | 1.000 |
| Thiamine Metabolism | Cofactors and Vitamins | 15.974 | 8.300 | 0.117 | 1.000 |
| Fatty Acid Metabolism (also BCAA Metabolism) | Lipid | 7.974 | 5.533 | 0.170 | 1.000 |
| Guanidino and Acetamido Metabolism | Amino Acid | 7.974 | 5.533 | 0.170 | 1.000 |
| Urea cycle; Arginine and Proline Metabolism | Amino Acid | 2.119 | 1.915 | 0.202 | 1.000 |
| Tyrosine Metabolism | Amino Acid | 0.000 | 0.000 | 0.248 | 1.000 |
| Fatty Acid Metabolism (Acyl Carnitine, Medium Chain) | Lipid | 3.974 | 3.320 | 0.268 | 1.000 |
| Disaccharides and Oligosaccharides | Carbohydrate | 2.641 | 2.371 | 0.354 | 1.000 |
| Pregnenolone Steroids | Lipid | 2.641 | 2.371 | 0.354 | 1.000 |
| Fatty Acid, Dicarboxylate | Lipid | 1.676 | 1.581 | 0.365 | 1.000 |
| Food Component/Plant | Xenobiotics | 1.500 | 1.423 | 0.461 | 1.000 |
| Androgenic Steroids | Lipid | 1.752 | 1.660 | 0.465 | 1.000 |
| Fatty Acid Metabolism (Acyl Glycine) | Lipid | 1.752 | 1.660 | 0.465 | 1.000 |
| TCA Cycle | Energy | 1.205 | 1.186 | 0.585 | 1.000 |
| Pyrimidine Metabolism, Uracil containing | Nucleotide | 1.117 | 1.107 | 0.610 | 1.000 |
| Tryptophan Metabolism | Amino Acid | 0.000 | 0.000 | 0.631 | 1.000 |
| Benzoate Metabolism | Xenobiotics | 0.000 | 0.000 | 0.636 | 1.000 |
| Chemical | Xenobiotics | 0.000 | 0.000 | 0.636 | 1.000 |
| Methionine, Cysteine, SAM and Taurine Metabolism | Amino Acid | 0.000 | 0.000 | 0.636 | 1.000 |
| Histidine Metabolism | Amino Acid | 1.511 | 1.443 | 0.643 | 1.000 |
| Leucine, Isoleucine and Valine Metabolism | Amino Acid | 1.120 | 1.107 | 0.701 | 1.000 |
| Acetylated Peptides | Peptide | 0.000 | 0.000 | 1.000 | 1.000 |
| Advanced Glycation End-product | Carbohydrate | 0.000 | 0.000 | 1.000 | 1.000 |
| Alanine and Aspartate Metabolism | Amino Acid | 0.000 | 0.000 | 1.000 | 1.000 |
| Aminosugar Metabolism | Carbohydrate | 0.000 | 0.000 | 1.000 | 1.000 |
| Bacterial/Fungal | Xenobiotics | 0.000 | 0.000 | 1.000 | 1.000 |
| Carnitine Metabolism | Lipid | 0.000 | 0.000 | 1.000 | 1.000 |
| Corticosteroids | Lipid | 0.000 | 0.000 | 1.000 | 1.000 |
| Creatine Metabolism | Amino Acid | 0.000 | 0.000 | 1.000 | 1.000 |
| Dipeptide | Peptide | 0.000 | 0.000 | 1.000 | 1.000 |
| Dipeptide Derivative | Peptide | 0.000 | 0.000 | 1.000 | 1.000 |

| | | | | | |
|---|---|---|---|---|---|
| Drug - Analgesics, Anesthetics | Xenobiotics | 0.000 | 0.000 | 1.000 | 1.000 |
| Drug - Topical Agents | Xenobiotics | 0.000 | 0.000 | 1.000 | 1.000 |
| Fatty Acid Metabolism (Acyl Glutamine) | Lipid | 0.000 | 0.000 | 1.000 | 1.000 |
| Fatty Acid, Branched | Lipid | 0.000 | 0.000 | 1.000 | 1.000 |
| Fatty Acid, Dihydroxy | Lipid | 0.000 | 0.000 | 1.000 | 1.000 |
| Fatty Acid, Monohydroxy | Lipid | 0.000 | 0.000 | 1.000 | 1.000 |
| Fructose, Mannose and Galactose Metabolism | Carbohydrate | 0.000 | 0.000 | 1.000 | 1.000 |
| Gamma-glutamyl Amino Acid | Peptide | 0.000 | 0.000 | 1.000 | 1.000 |
| Glutamate Metabolism | Amino Acid | 0.000 | 0.000 | 1.000 | 1.000 |
| Glutathione Metabolism | Amino Acid | 0.000 | 0.000 | 1.000 | 1.000 |
| Glycerolipid Metabolism | Lipid | 0.000 | 0.000 | 1.000 | 1.000 |
| Glycine, Serine and Threonine Metabolism | Amino Acid | 0.000 | 0.000 | 1.000 | 1.000 |
| Glycolysis, Gluconeogenesis, and Pyruvate Metabolism | Carbohydrate | 0.000 | 0.000 | 1.000 | 1.000 |
| Lactoyl Amino Acid | Amino Acid | 0.000 | 0.000 | 1.000 | 1.000 |
| Long Chain Monounsaturated Fatty Acid | Lipid | 0.000 | 0.000 | 1.000 | 1.000 |
| Long Chain Polyunsaturated Fatty Acid (n3 and n6) | Lipid | 0.000 | 0.000 | 1.000 | 1.000 |
| Long Chain Saturated Fatty Acid | Lipid | 0.000 | 0.000 | 1.000 | 1.000 |
| Mevalonate Metabolism | Lipid | 0.000 | 0.000 | 1.000 | 1.000 |
| Nicotinate and Nicotinamide Metabolism | Cofactors and Vitamins | 0.000 | 0.000 | 1.000 | 1.000 |
| Pantothenate and CoA Metabolism | Cofactors and Vitamins | 0.000 | 0.000 | 1.000 | 1.000 |
| Pentose Metabolism | Carbohydrate | 0.000 | 0.000 | 1.000 | 1.000 |
| Phenylalanine Metabolism | Amino Acid | 0.000 | 0.000 | 1.000 | 1.000 |
| Phospholipid Metabolism | Lipid | 0.000 | 0.000 | 1.000 | 1.000 |
| Polyamine Metabolism | Amino Acid | 0.000 | 0.000 | 1.000 | 1.000 |
| Primary Bile Acid Metabolism | Lipid | 0.000 | 0.000 | 1.000 | 1.000 |
| Progestin Steroids | Lipid | 0.000 | 0.000 | 1.000 | 1.000 |
| Pterin Metabolism | Cofactors and Vitamins | 0.000 | 0.000 | 1.000 | 1.000 |
| Purine Metabolism, Adenine containing | Nucleotide | 0.000 | 0.000 | 1.000 | 1.000 |
| Purine Metabolism, Guanine containing | Nucleotide | 0.000 | 0.000 | 1.000 | 1.000 |
| Pyrimidine Metabolism, Cytidine containing | Nucleotide | 0.000 | 0.000 | 1.000 | 1.000 |
| Pyrimidine Metabolism, Orotate containing | Nucleotide | 0.000 | 0.000 | 1.000 | 1.000 |
| Pyrimidine Metabolism, Thymine containing | Nucleotide | 0.000 | 0.000 | 1.000 | 1.000 |
| Secondary Bile Acid Metabolism | Lipid | 0.000 | 0.000 | 1.000 | 1.000 |
| Tetrahydrobiopterin Metabolism | Cofactors and Vitamins | 0.000 | 0.000 | 1.000 | 1.000 |
| Tocopherol Metabolism | Cofactors and Vitamins | 0.000 | 0.000 | 1.000 | 1.000 |
| Xanthine Metabolism | Xenobiotics | 0.000 | 0.000 | 1.000 | 1.000 |

*p* p-value; *q* q-value

An enrichment ratio >1 indicates overrepresentation of BPD-differential metabolites within the sub-pathway.

**Table E15: Summary statistics of sub-pathway enrichment analysis at time point 2 (postnatal days 23-32) for infants on enteral nutrition**

| Sub-Pathway | Super-Pathway | Odds Ratio | Enrichment Ratio | *p* | *q* |
|---|---|---|---|---|---|
| Tryptophan Metabolism | Amino Acid | 5.192 | 4.124 | 0.032 | 1.000 |
| Androgenic Steroids | Lipid | 7.536 | 5.774 | 0.044 | 1.000 |
| Polyamine Metabolism | Amino Acid | 6.010 | 4.812 | 0.061 | 1.000 |
| Dipeptide Derivative | Peptide | 29.091 | 14.435 | 0.068 | 1.000 |
| Mevalonate Metabolism | Lipid | 14.523 | 9.623 | 0.101 | 1.000 |
| Glutathione Metabolism | Amino Acid | 9.667 | 7.217 | 0.132 | 1.000 |
| Phenylalanine Metabolism | Amino Acid | 4.811 | 4.124 | 0.220 | 1.000 |
| Aminosugar Metabolism | Carbohydrate | 4.117 | 3.609 | 0.247 | 1.000 |
| Ascorbate and Aldarate Metabolism | Cofactors and Vitamins | 4.117 | 3.609 | 0.247 | 1.000 |
| Dipeptide | Peptide | 3.597 | 3.208 | 0.273 | 1.000 |
| Xanthine Metabolism | Xenobiotics | 2.603 | 2.406 | 0.347 | 1.000 |
| TCA Cycle | Energy | 2.196 | 2.062 | 0.393 | 1.000 |
| Chemical | Xenobiotics | 1.342 | 1.312 | 0.545 | 1.000 |
| Methionine, Cysteine, SAM and Taurine Metabolism | Amino Acid | 1.342 | 1.312 | 0.545 | 1.000 |
| Lysine Metabolism | Amino Acid | 1.221 | 1.203 | 0.577 | 1.000 |
| Urea cycle; Arginine and Proline Metabolism | Amino Acid | 1.120 | 1.110 | 0.607 | 1.000 |
| Food Component/Plant | Xenobiotics | 0.000 | 0.000 | 0.627 | 1.000 |
| Acetylated Peptides | Peptide | 0.000 | 0.000 | 1.000 | 1.000 |
| Advanced Glycation End-product | Carbohydrate | 0.000 | 0.000 | 1.000 | 1.000 |
| Alanine and Aspartate Metabolism | Amino Acid | 0.000 | 0.000 | 1.000 | 1.000 |
| Bacterial/Fungal | Xenobiotics | 0.000 | 0.000 | 1.000 | 1.000 |
| Benzoate Metabolism | Xenobiotics | 0.000 | 0.000 | 1.000 | 1.000 |
| Carnitine Metabolism | Lipid | 0.000 | 0.000 | 1.000 | 1.000 |
| Corticosteroids | Lipid | 0.000 | 0.000 | 1.000 | 1.000 |
| Creatine Metabolism | Amino Acid | 0.000 | 0.000 | 1.000 | 1.000 |
| Disaccharides and Oligosaccharides | Carbohydrate | 0.000 | 0.000 | 1.000 | 1.000 |
| Drug - Analgesics, Anesthetics | Xenobiotics | 0.000 | 0.000 | 1.000 | 1.000 |
| Drug - Topical Agents | Xenobiotics | 0.000 | 0.000 | 1.000 | 1.000 |
| Fatty Acid Metabolism (Acyl Carnitine, Dicarboxylate) | Lipid | 0.000 | 0.000 | 1.000 | 1.000 |
| Fatty Acid Metabolism (Acyl Carnitine, Hydroxy) | Lipid | 0.000 | 0.000 | 1.000 | 1.000 |
| Fatty Acid Metabolism (Acyl Carnitine, Medium Chain) | Lipid | 0.000 | 0.000 | 1.000 | 1.000 |
| Fatty Acid Metabolism (Acyl Glutamine) | Lipid | 0.000 | 0.000 | 1.000 | 1.000 |
| Fatty Acid Metabolism (Acyl Glycine) | Lipid | 0.000 | 0.000 | 1.000 | 1.000 |
| Fatty Acid Metabolism (also BCAA Metabolism) | Lipid | 0.000 | 0.000 | 1.000 | 1.000 |
| Fatty Acid Synthesis | Lipid | 0.000 | 0.000 | 1.000 | 1.000 |
| Fatty Acid, Branched | Lipid | 0.000 | 0.000 | 1.000 | 1.000 |
| Fatty Acid, Dicarboxylate | Lipid | 0.000 | 0.000 | 1.000 | 1.000 |

| | | | | | |
|---|---|---|---|---|---|
| Fatty Acid, Dihydroxy | Lipid | 0.000 | 0.000 | 1.000 | 1.000 |
| Fatty Acid, Monohydroxy | Lipid | 0.000 | 0.000 | 1.000 | 1.000 |
| Fructose, Mannose and Galactose Metabolism | Carbohydrate | 0.000 | 0.000 | 1.000 | 1.000 |
| Gamma-glutamyl Amino Acid | Peptide | 0.000 | 0.000 | 1.000 | 1.000 |
| Glutamate Metabolism | Amino Acid | 0.000 | 0.000 | 1.000 | 1.000 |
| Glycerolipid Metabolism | Lipid | 0.000 | 0.000 | 1.000 | 1.000 |
| Glycine, Serine and Threonine Metabolism | Amino Acid | 0.000 | 0.000 | 1.000 | 1.000 |
| Glycolysis, Gluconeogenesis, and Pyruvate Metabolism | Carbohydrate | 0.000 | 0.000 | 1.000 | 1.000 |
| Guanidino and Acetamido Metabolism | Amino Acid | 0.000 | 0.000 | 1.000 | 1.000 |
| Histidine Metabolism | Amino Acid | 0.000 | 0.000 | 1.000 | 1.000 |
| Lactoyl Amino Acid | Amino Acid | 0.000 | 0.000 | 1.000 | 1.000 |
| Leucine, Isoleucine and Valine Metabolism | Amino Acid | 0.959 | 0.962 | 1.000 | 1.000 |
| Long Chain Monounsaturated Fatty Acid | Lipid | 0.000 | 0.000 | 1.000 | 1.000 |
| Long Chain Polyunsaturated Fatty Acid (n3 and n6) | Lipid | 0.000 | 0.000 | 1.000 | 1.000 |
| Long Chain Saturated Fatty Acid | Lipid | 0.000 | 0.000 | 1.000 | 1.000 |
| Nicotinate and Nicotinamide Metabolism | Cofactors and Vitamins | 0.000 | 0.000 | 1.000 | 1.000 |
| Pantothenate and CoA Metabolism | Cofactors and Vitamins | 0.000 | 0.000 | 1.000 | 1.000 |
| Pentose Metabolism | Carbohydrate | 0.000 | 0.000 | 1.000 | 1.000 |
| Phospholipid Metabolism | Lipid | 0.000 | 0.000 | 1.000 | 1.000 |
| Pregnenolone Steroids | Lipid | 0.000 | 0.000 | 1.000 | 1.000 |
| Primary Bile Acid Metabolism | Lipid | 0.000 | 0.000 | 1.000 | 1.000 |
| Progestin Steroids | Lipid | 0.000 | 0.000 | 1.000 | 1.000 |
| Pterin Metabolism | Cofactors and Vitamins | 0.000 | 0.000 | 1.000 | 1.000 |
| Purine Metabolism, (Hypo)Xanthine/Inosine containing | Nucleotide | 0.000 | 0.000 | 1.000 | 1.000 |
| Purine Metabolism, Adenine containing | Nucleotide | 0.000 | 0.000 | 1.000 | 1.000 |
| Purine Metabolism, Guanine containing | Nucleotide | 0.000 | 0.000 | 1.000 | 1.000 |
| Pyrimidine Metabolism, Cytidine containing | Nucleotide | 0.000 | 0.000 | 1.000 | 1.000 |
| Pyrimidine Metabolism, Orotate containing | Nucleotide | 0.000 | 0.000 | 1.000 | 1.000 |
| Pyrimidine Metabolism, Thymine containing | Nucleotide | 0.000 | 0.000 | 1.000 | 1.000 |
| Pyrimidine Metabolism, Uracil containing | Nucleotide | 0.000 | 0.000 | 1.000 | 1.000 |
| Secondary Bile Acid Metabolism | Lipid | 0.000 | 0.000 | 1.000 | 1.000 |
| Tetrahydrobiopterin Metabolism | Cofactors and Vitamins | 0.000 | 0.000 | 1.000 | 1.000 |
| Thiamine Metabolism | Cofactors and Vitamins | 0.000 | 0.000 | 1.000 | 1.000 |
| Tocopherol Metabolism | Cofactors and Vitamins | 0.000 | 0.000 | 1.000 | 1.000 |
| Tyrosine Metabolism | Amino Acid | 0.959 | 0.962 | 1.000 | 1.000 |
| Vitamin B6 Metabolism | Cofactors and Vitamins | 0.000 | 0.000 | 1.000 | 1.000 |

*p* p-value; *q* q-value

An enrichment ratio >1 indicates overrepresentation of BPD-differential metabolites within the sub-pathway.
